# A Multi-Country Evaluation of Patient Preferences for Tuberculosis Diagnostics: A Discrete Choice Experiment to Inform WHO Target Product Profiles

**DOI:** 10.1101/2024.06.19.24309124

**Authors:** Kinari Shah, Talemwa Nalugwa, Danaida Marcelo, Robinah Nakawunde, Trang Trinh, Deepa Shankar, Annet Nakaweesa, Ann Schraufnagel, Alfred Andama, DJ Christopher, Dinh Van Luong, Grant Theron, William Worodria, Maria del Mar Castro, Payam Nahid, Claudia M. Denkinger, Adithya Cattamanchi, Charles Yu, Andrew D. Kerkhoff

**Affiliations:** Division of Pulmonary Diseases and Critical Care Medicine, University of California San Francisco, San Francisco, CA USA; Center for Tuberculosis, University of California San Francisco, San Francisco, CA USA; World Alliance for Lung and Intensive Care Medicines in Uganda (WALIMU), Kampala, Uganda; Research Services, De La Salle Medical and Health Sciences Institute, Cavite, Philippines; Division of Molecular Biology and Human Genetics, Stellenbosch University, Cape Town South Africa; Vietnam National Tuberculosis Program, University of California San Francisco Research Collaboration Unit; Center for Promotion of Advancement of Society, Hanoi, Vietnam; Department of Pulmonary Medicine, Christian Medical College, Vellore, Tamil Nadu, India; National Lung Hospital, Hanoi, Vietnam; Department of Infectious Disease, German Centre for Infection Research (DZIF), Partner site Heidelberg University Hospital and Faculty of Medicine, Heidelberg, Germany; Institute for Global Health Sciences, University of California San Francisco, San Francisco, CA USA; University of California Irvine, Division of Pulmonary Diseases and Critical Care Medicine, Irvine, United States of America; Division of HIV, Infectious Diseases, and Global Medicine, University of California San Francisco, San Francisco, CA USA

**Author notes:** Corresponding author: Kinari Shah. Co-first authors (Kinari Shah, Talemwa Nalugwa). Senior author (Charles Yu, Andrew D. Kerkhoff).

## Abstract

**Background:** Recognizing new and improved diagnostics as a key step to reducing the global tuberculosis (TB) burden, the World Health Organization (WHO) publishes target product profiles (TPPs) to guide development of novel TB diagnostics; however, TPPs have not included the preferences of people undergoing TB testing. Understanding their preferences is crucial for optimizing the acceptability and uptake of TB diagnostic services.

**Design/methods:** We conducted a discrete choice experiment (DCE) among adults with presumptive or microbiologically confirmed TB in 5 countries (Philippines, Vietnam, South Africa, Uganda, and India). The DCE evaluated preferences for 5 TB diagnostic test attributes (sample type, accuracy, cost, location, time to result) with 3-4 levels per attribute. We estimated mean preference weights for attribute levels using Hierarchical Bayesian models and conducted willingness-to-trade analyses for preferred test features.

**Results:** Among 1,033 participants (median age 43 years, Interquartile Range: 30-55), 52% were female, 10% had HIV, 14% had diabetes, 36% had current or prior TB), the most strongly valued test features were free cost, high diagnostic accuracy, and rapid (within 15 minutes) result availability. Among less valued attributes, facility-based testing (location) and sputum (sample type) were most preferred. The relative importance of attributes and their levels differed by country. In willingness-to-trade simulations, participants in each country were willing to trade 10-20% lower accuracy for the availability of rapid tests but were unwilling to trade accuracy for alternative sample types (e.g., tongue-swab, urine, blood) or testing locations (e.g., home- or community-based); participants in India and Vietnam were willing to pay $3.20 and $4.00 USD, respectively, for tests with rapid results.

**Conclusions:** Persons accessing TB services in five high-burden countries universally value free cost, high diagnostic accuracy, and rapid result availability. However, many were willing to accept lower test accuracy and some were willing to incur small out-of-pocket costs to access rapid tests. To align with preferences of TB-affected communities, future TPP updates and new test development should prioritize rapid results while maintaining high accuracy and minimizing costs.

## Introduction

With more than 10 million new cases and 1.3 million attributable deaths each year, tuberculosis (TB) remains a significant global public health challenge^1^. Missed diagnoses account for the largest gap in the global TB care cascade and represent a major barrier to improved control^1^. People with undiagnosed TB living in high-burden settings may face substantial challenges at each step of their care journey; initiating healthcare seeking for their symptoms, accessing TB testing services, receiving a diagnosis and starting on treatment^2^. To close gaps along this care cascade, there is an important need to better understand perspectives and preferences of TB affected communities in order to better align TB tools and services with their needs and wants.

While there is increasing focus on offering ‘people-centered’ TB services^3,4^, there has been little attention to whether current TB diagnostic tools are acceptable to people affected by TB and if future TB diagnostic tools and services could be more appealing and would promote engagement if they contained their preferred features. The World Health Organization Target product profiles (TPPs) for new TB diagnostics outline the minimal and ideal features and accuracy thresholds needed for new TB tests^5,6^. They include the views of government officials, providers, and implementing partners; however, notably, the views of those undergoing TB testing have not been directly considered or included. This gap highlights the need for more inclusive research that aims to understand and prioritize the preferences of those directly affected by TB.

Aligning diagnostic tools with preferences of people with TB could significantly enhance the effectiveness of TB control efforts by improving acceptability and access, leading to better TB detection rates and timely treatment initiation. Preference methods, and in particular discrete choice experiments (DCEs), have the potential to provide valuable insights into what people with TB prefer and value most in diagnostic tools^7^. Challenges and considerations such as the invasiveness of sample collection, test accuracy, the time required for test results, and the convenience and cost of testing are all critical factors that can influence engagement in TB diagnostic services. Since it may not be possible to optimize all features simultaneously in a single diagnostic test, understanding the trade-offs that people are willing to make is crucial. DCEs can help to characterize these trade-offs, thereby informing the development of more person-centered diagnostic tools.

In order to better understand preferences of people with presumed or confirmed TB for TB diagnostic tests and inform future TPP revisions, we conducted a DCE in five high TB burden countries. Our objectives were to determine the relative importance of different TB test features and to assess whether participants were willing to trade lower test accuracy for more desirable test features.

## Methods

### Setting and Participants

A cross-sectional survey was administered at outpatient health centers located in five countries: Vellore, India; Dasmarinas, Philippines; Cape Town, South Africa; Hanoi, Vietnam; and Kampala, Uganda, between November 2022 and May 2024. Eligibility criteria included people ≥18 years who met country-specific presumptive TB definitions or had undergone TB testing within the three months preceding screening for study inclusion and who provided informed consent. People cognitively unable to complete DCE choice tasks (e.g., too sick) were excluded. A detailed description of the study setting and population by country can be found in **Table S1**.

### Ethics, consent, and permissions

Institutional approval was obtained from De La Salle Medical Health & Sciences Institute in the Philippines, Hanoi Lung Hospital in Vietnam, Stellenbosch University in South Africa, Makerere University in Uganda, Christian Medical College in India, University of Heidelberg in Germany, and the University of California San Francisco. All participants provided written or verbal informed consent in their preferred language.

### Study Design and Procedures

The DCE attributes and levels were selected after a review of the literature and TPP requirements and through discussions with implementing partners and a research committee including other co-investigators. Initially, six attributes with up to four attribute levels were selected for the DCE design, which included both specificity and sensitivity as separate attributes. However, after discussions with partners, due to the complexity of explaining specificity and concerns about its comprehension, we removed it from the design. Instead, only a single, simplified test accuracy attribute, representative of sensitivity was included. Thus, the final DCE had 5 attributes with 3-4 levels each (**Figure 1**). To minimize potential issues related to limited health literacy, standardized graphics were developed to represent each test feature (**Figure 1**). Additionally, in each country, the cost attribute was adjusted to account for the relative equivalent cost in the local currency^8^.

**Figure 1.**
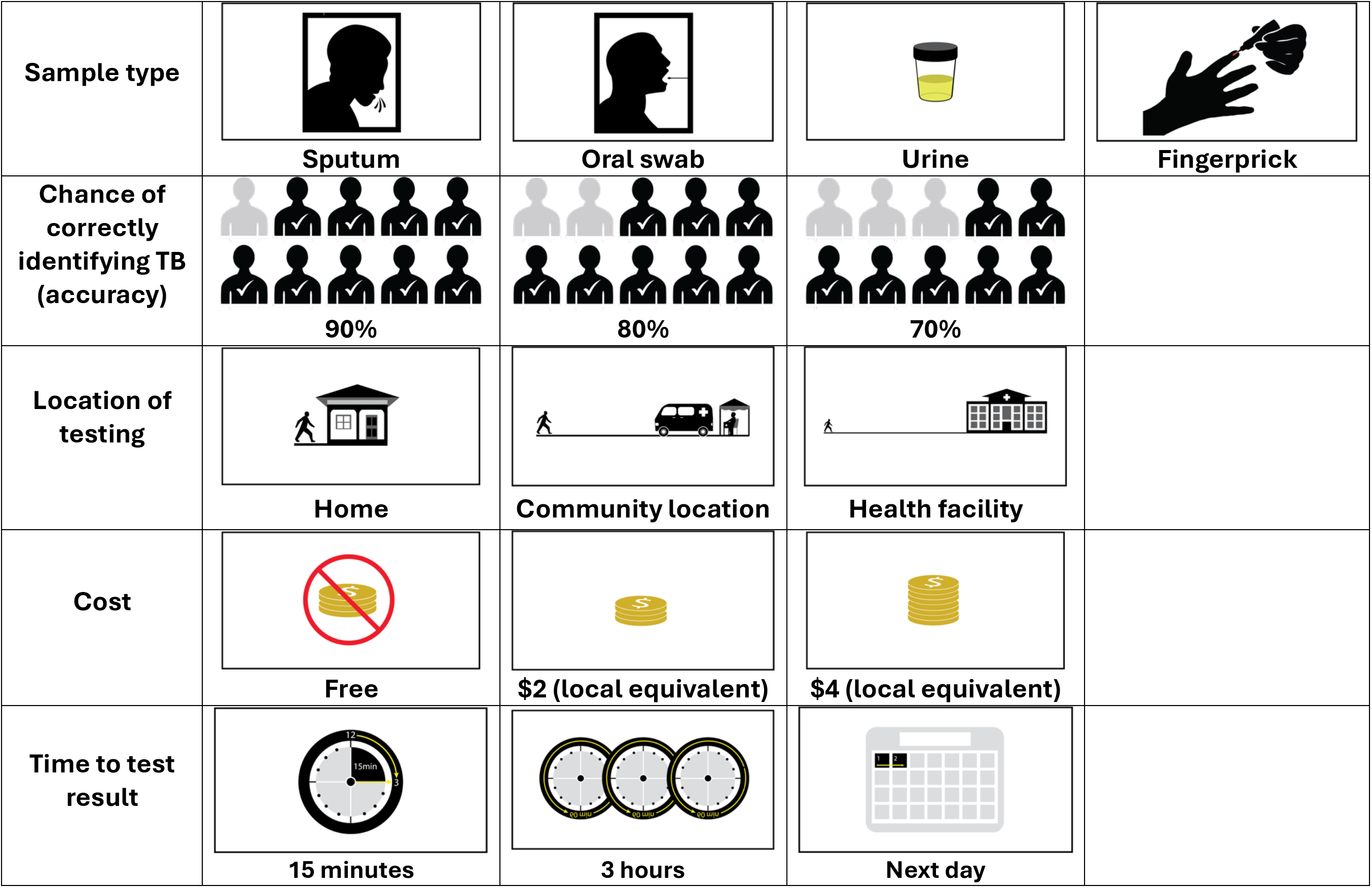
Final DCE attributes and levels including figures displayed to each respondent.

During the design of the DCE, research staff from each country reviewed the survey, training materials, and graphics and provided detailed feedback on all participant materials and study procedures. This included a participant education handbook, which was utilized to facilitate a standardized understanding of the DCE test features across diverse settings (**Appendix 1**). Each country team also provided translations for the handbook and DCE to ensure participants completed the exercise in their preferred language. Across all sites, props were utilized to better demonstrate sputum, oral swab, finger blood prick, and urine sample obtainment. This was done to ensure that hypothetical bias was minimized for participants who may not have previously experienced the collection of all sample types^9^.

To assess for the need refinements to the study design and procedures and to standardize participant education and survey administration, each site completed a pilot of the DCE procedures among 5-10 participants. During this process, each country site assessed the adequacy of the participant education materials and translations in facilitating understanding of the graphics and choice tasks, as well as participant comprehension of the overall exercise and choice tasks. The piloting did not result in major changes to the DCE design or attributes/levels but led to minor adjustments in translated explanations to improve local understanding.

The final DCE was designed in Sawtooth Software version 9.14.2 and used a balanced overlap design that was nearly level-balanced and orthogonal. After reviewing the participant education handbook and completing a fixed “warm-up task,” participants were shown 12 random choice tasks; this number was selected to minimize cognitive burden, while ensuring that each level was seen six times to allow for adequate precision in individual-level preference estimates (Figure S1)^10,11^. In each choice task, participants were asked to choose one of two hypothetical TB diagnostic tests that they most preferred based on their review of the distinct attribute levels for each test. After making a choice, participants were then asked whether they would get tested with their preferred test (i.e., dual-response none design). To assess comprehension and DCE engagement, participants were also shown one fixed, dominant choice task featuring a hypothetical TB test that was clearly better than the other option. After completing all choice tasks, participants were shown three Likert-scale questions to assess their understanding of the choice tasks, their ability to differentiate between the test options shown, and the difficulty of choosing between the test options.

Eligible participants were consecutively enrolled at each health facility. Upon enrollment, participants completed a structured demographics survey and the DCE with the help of a trained research team member using an electronic tablet with Sawtooth offline surveys.

### Sample Size

The sample size calculation for a DCE is based on the formula: *500*I/(J*S)*, where I is the product of the two attributes with the highest levels, J is the number of alternatives presented in a choice task, and S is the total number of choice tasks. Based on the design, the minimum sample size for this DCE was n=313 [500(5*3)/(2*12)]. However, a minimum sample size of DCE is recommended per sub-group to generate meaningful estimates. Therefore, the total sample size per country was set at 200 respondents meeting data quality standards, for a total of 1000 included participants^10^. The quality of DCE responses was monitored to ensure that each country met its target.

## Statistical Analysis

### Data quality

Prior to the analysis, we applied several measures to ensure high-quality individual-level responses. First, we excluded all participants who failed the dominant choice task or always selected test option A or test option B in every choice tasks (i.e., straightlining)^12^. We also excluded those who met two of the three following criteria: 1) self-reported difficulty with understanding choice tasks or differentiating between the two test options, 2) completed the DCE in under 40% of median country-specific response time, 3) did not exceed the 95% internal consistency fit statistic threshold indicating a likely inattentive or ‘random responder;’ this value was generated using 1000 bots completing the DCE at random^13,14^.

### Analyses

Analyses were conducted using Lighthouse Studio version 14.1.2 (Sawtooth Software) and Stata 17.0 (StataCorp LLC). Hierarchical Bayesian (HB) models were used to calculate zero-centered individual-level and overall mean preference weights (MPW) (i.e., part-worth utilities) of attribute levels and relative importance (RI) of attributes, overall and by country. MPW were derived by averaging 10,000 iterative estimations of parameters based on each respondent’s data.

The primary analysis focused on differences in MPW and RI by country, and also key demographic variables, including gender, self-reported diabetes status, self-reported HIV status, prior TB disease status, and distance traveled to the facility. Using HB-derived individual-level MPW, we also conducted shares of preference simulations in the Sawtooth Simulator to estimate what amount of test accuracy participants would be willing to trade for preferred test features and what amount participants were willing to pay for specific test features^11^. Sensitivity analyses were undertaken to assess the impact of excluding participants who did not meet data quality standards.

## Results

### Participant characteristics

Of 1,130 participants enrolled across all countries we excluded 97 (8.6%) who did not meet the quality standards. Among the remaining 1,033 participants included in the final analysis, 200 (19.4%) were from India, 210 (20.3%) from the Philippines, 216 (20.9%) from South Africa, 200 (19.4%) from Uganda, and 208 (20.1%) from Vietnam. Overall, the median age was 43 years (IQR 30-55), 533 (52%) were female, 105 (10%) were HIV-positive, 149 (14%) were diabetic, and 364 (36%) had current or prior TB (**Table 1**). Participants in Uganda and South Africa tended to be younger and were more likely to be female and HIV-positive, while in India and the Philippines, there was a higher proportion of diabetic participants; participants in India and Vietnam traveled much further to access TB services. Notably, the characteristics of participants excluded due to quality checks did not meaningfully differ from those included.

**Table 1.**
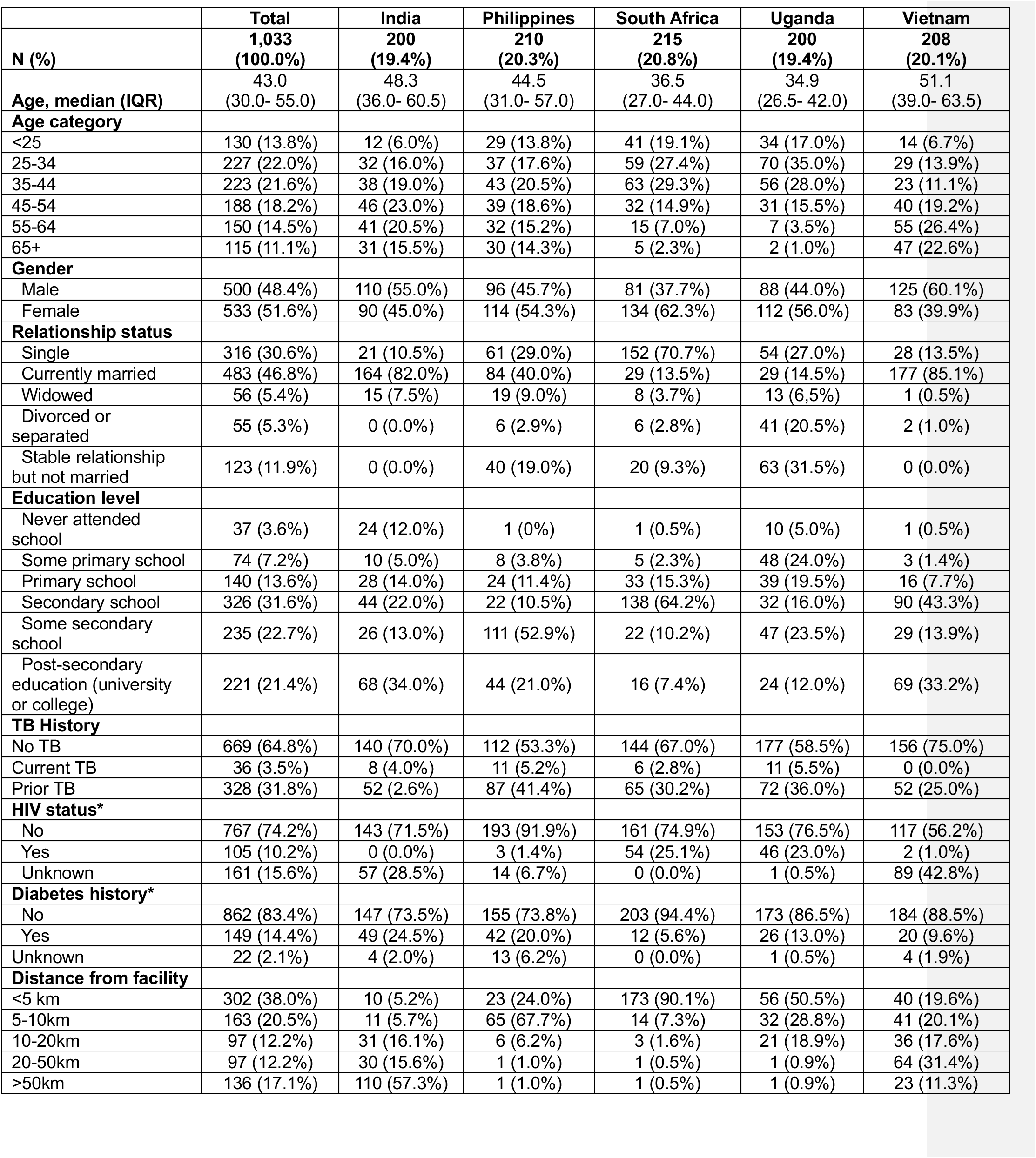
Participant characteristics by country.

|  | **Total** | **India** | **Philippines** | **South Africa** | **Uganda** | **Vietnam** |
| --- | --- | --- | --- | --- | --- | --- |
| **N (%)** | **1,033**<br>**(100.0%)** | **200**<br>**(19.4%)** | **210**<br>**(20.3%)** | **215**<br>**(20.8%)** | **200**<br>**(19.4%)** | **208**<br>**(20.1%)** |
| **Age, median (IQR)** | 43.0<br>(30.0- 55.0) | 48.3<br>(36.0- 60.5) | 44.5<br>(31.0- 57.0) | 36.5<br>(27.0- 44.0) | 34.9<br>(26.5- 42.0) | 51.1<br>(39.0- 63.5) |
| **Age category** |  |  |  |  |  |  |
| <25 | 130 (13.8%) | 12 (6.0%) | 29 (13.8%) | 41 (19.1%) | 34 (17.0%) | 14 (6.7%) |
| 25-34 | 227 (22.0%) | 32 (16.0%) | 37 (17.6%) | 59 (27.4%) | 70 (35.0%) | 29 (13.9%) |
| 35-44 | 223 (21.6%) | 38 (19.0%) | 43 (20.5%) | 63 (29.3%) | 56 (28.0%) | 23 (11.1%) |
| 45-54 | 188 (18.2%) | 46 (23.0%) | 39 (18.6%) | 32 (14.9%) | 31 (15.5%) | 40 (19.2%) |
| 55-64 | 150 (14.5%) | 41 (20.5%) | 32 (15.2%) | 15 (7.0%) | 7 (3.5%) | 55 (26.4%) |
| 65+ | 115 (11.1%) | 31 (15.5%) | 30 (14.3%) | 5 (2.3%) | 2 (1.0%) | 47 (22.6%) |
| **Gender** |  |  |  |  |  |  |
| Male | 500 (48.4%) | 110 (55.0%) | 96 (45.7%) | 81 (37.7%) | 88 (44.0%) | 125 (60.1%) |
| Female | 533 (51.6%) | 90 (45.0%) | 114 (54.3%) | 134 (62.3%) | 112 (56.0%) | 83 (39.9%) |
| **Relationship status** |  |  |  |  |  |  |
| Single | 316 (30.6%) | 21 (10.5%) | 61 (29.0%) | 152 (70.7%) | 54 (27.0%) | 28 (13.5%) |
| Currently married | 483 (46.8%) | 164 (82.0%) | 84 (40.0%) | 29 (13.5%) | 29 (14.5%) | 177 (85.1%) |
| Widowed | 56 (5.4%) | 15 (7.5%) | 19 (9.0%) | 8 (3.7%) | 13 (6.5%) | 1 (0.5%) |
| Divorced or separated | 55 (5.3%) | 0 (0.0%) | 6 (2.9%) | 6 (2.8%) | 41 (20.5%) | 2 (1.0%) |
| Stable relationship but not married | 123 (11.9%) | 0 (0.0%) | 40 (19.0%) | 20 (9.3%) | 63 (31.5%) | 0 (0.0%) |
| **Education level** |  |  |  |  |  |  |
| Never attended school | 37 (3.6%) | 24 (12.0%) | 1 (0%) | 1 (0.5%) | 10 (5.0%) | 1 (0.5%) |
| Some primary school | 74 (7.2%) | 10 (5.0%) | 8 (3.8%) | 5 (2.3%) | 48 (24.0%) | 3 (1.4%) |
| Primary school | 140 (13.6%) | 28 (14.0%) | 24 (11.4%) | 33 (15.3%) | 39 (19.5%) | 16 (7.7%) |
| Secondary school | 326 (31.6%) | 44 (22.0%) | 22 (10.5%) | 138 (64.2%) | 32 (16.0%) | 90 (43.3%) |
| Some secondary school | 235 (22.7%) | 26 (13.0%) | 111 (52.9%) | 22 (10.2%) | 47 (23.5%) | 29 (13.9%) |
| Post-secondary education (university or college) | 221 (21.4%) | 68 (34.0%) | 44 (21.0%) | 16 (7.4%) | 24 (12.0%) | 69 (33.2%) |
| **TB History** |  |  |  |  |  |  |
| No TB | 669 (64.8%) | 140 (70.0%) | 112 (53.3%) | 144 (67.0%) | 177 (58.5%) | 156 (75.0%) |
| Current TB | 36 (3.5%) | 8 (4.0%) | 11 (5.2%) | 6 (2.8%) | 11 (5.5%) | 0 (0.0%) |
| Prior TB | 328 (31.8%) | 52 (2.6%) | 87 (41.4%) | 65 (30.2%) | 72 (36.0%) | 52 (25.0%) |
| **HIV status\*** |  |  |  |  |  |  |
| No | 767 (74.2%) | 143 (71.5%) | 193 (91.9%) | 161 (74.9%) | 153 (76.5%) | 117 (56.2%) |
| Yes | 105 (10.2%) | 0 (0.0%) | 3 (1.4%) | 54 (25.1%) | 46 (23.0%) | 2 (1.0%) |
| Unknown | 161 (15.6%) | 57 (28.5%) | 14 (6.7%) | 0 (0.0%) | 1 (0.5%) | 89 (42.8%) |
| **Diabetes history\*** |  |  |  |  |  |  |
| No | 862 (83.4%) | 147 (73.5%) | 155 (73.8%) | 203 (94.4%) | 173 (86.5%) | 184 (88.5%) |
| Yes | 149 (14.4%) | 49 (24.5%) | 42 (20.0%) | 12 (5.6%) | 26 (13.0%) | 20 (9.6%) |
| Unknown | 22 (2.1%) | 4 (2.0%) | 13 (6.2%) | 0 (0.0%) | 1 (0.5%) | 4 (1.9%) |
| **Distance from facility** |  |  |  |  |  |  |
| <5 km | 302 (38.0%) | 10 (5.2%) | 23 (24.0%) | 173 (90.1%) | 56 (50.5%) | 40 (19.6%) |
| 5-10km | 163 (20.5%) | 11 (5.7%) | 65 (67.7%) | 14 (7.3%) | 32 (28.8%) | 41 (20.1%) |
| 10-20km | 97 (12.2%) | 31 (16.1%) | 6 (6.2%) | 3 (1.6%) | 21 (18.9%) | 36 (17.6%) |
| 20-50km | 97 (12.2%) | 30 (15.6%) | 1 (1.0%) | 1 (0.5%) | 1 (0.9%) | 64 (31.4%) |
| >50km | 136 (17.1%) | 110 (57.3%) | 1 (1.0%) | 1 (0.5%) | 1 (0.9%) | 23 (11.3%) |

### Preferences for TB test features overall and by country

Overall, the most important test attributes among all participants were cost (RI = 25.9), sensitivity (RI = 25.7), and time to test result (RI = 18.3), with free (MPW=59.7), highly (i.e., 90%) sensitive (MPW=57.3), and rapid (i.e., 15 minutes) tests (MPW=39.5), respectively, being the most preferred (**Figures 2 and 3).** Among the less valued attributes, for location of testing (RI=17.5), health facility-based testing was most preferred (MPW=21.0), while for sample type (RI=12.6) sputum was most preferred (MPW=4.1). While cost or sensitivity was the most important attribute in each country, several country-specific differences were observed regarding the relative importance of different test attributes, and the most preferred attribute levels (**Figures 2 and 3).** Notably, participants in India and the Philippines preferred tongue swabs to sputum samples, while participants in Vietnam preferred home-based testing.

**Figure 2.**
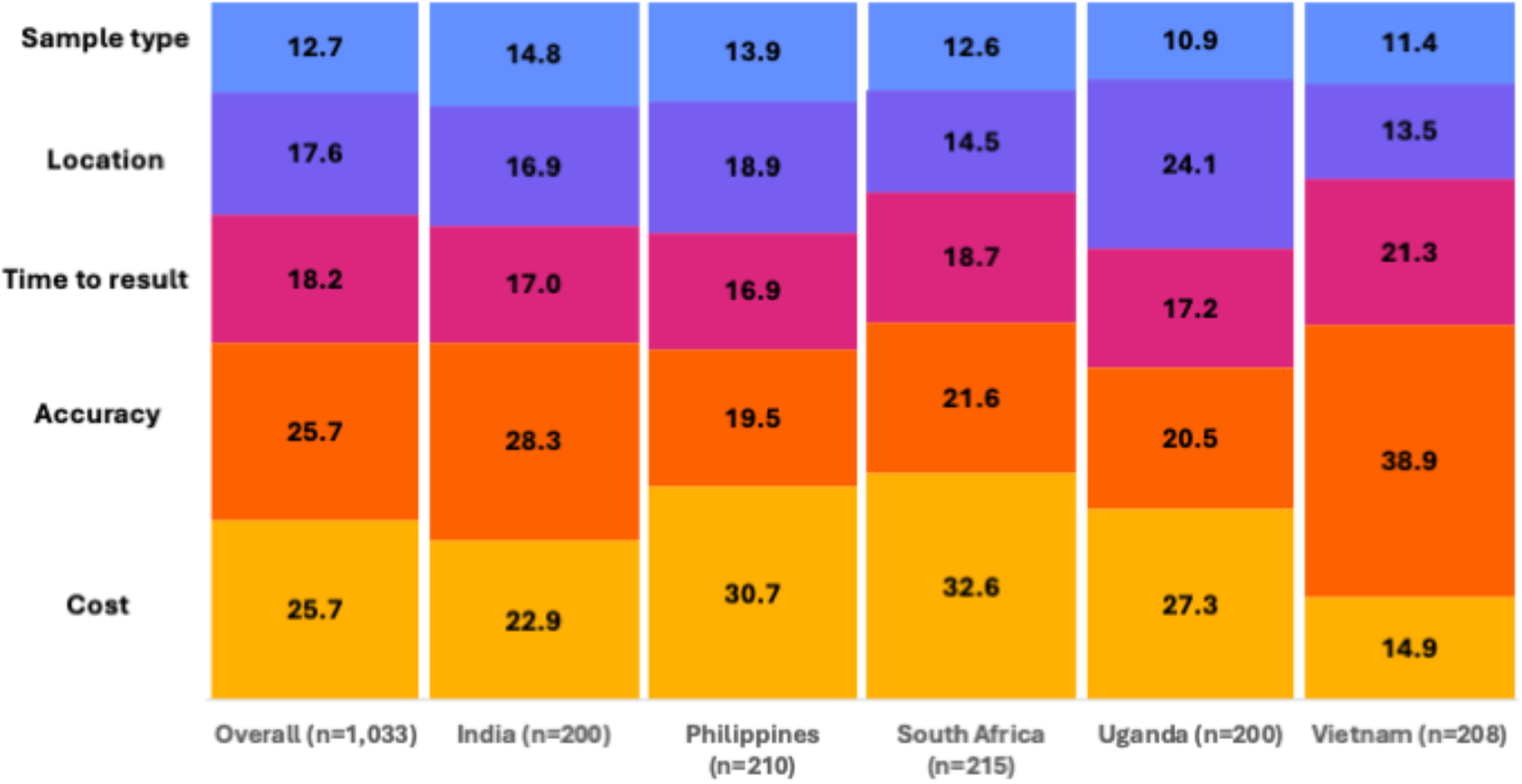
The relative importance of TB test attributes overall and by.

**Figure 3.**
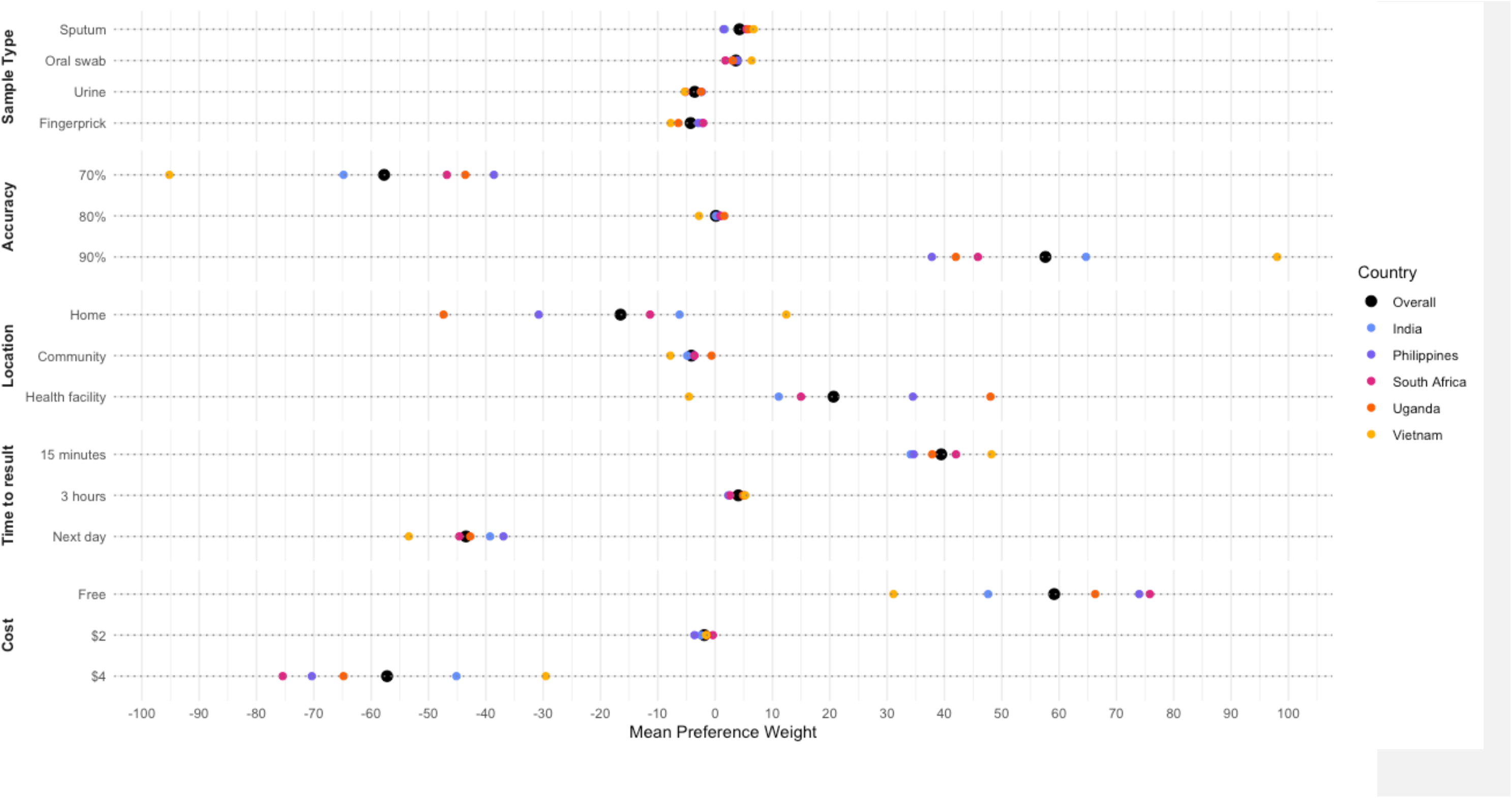
Mean Preference Weights for TB Test Features by Country.

Sensitivity analyses showed similar results when all participants were included, regardless of data quality (**Table S2**). Nearly all participants reported that the DCE was easy to understand (93.7%) and that it was easy to differentiate between the two options in each choice task (93.8%).

### Preferences for TB test characteristics by subgroup

Across all subgroups, highly accurate, free and rapid test results were the most important attributes and levels; sputum- and facility-based testing were also preferred among the less valued attributes across most subgroup analyses. However, participants with current or prior TB and those under 35 years of age preferred tongue swabs to sputum samples. Additionally, participants who traveled 20-50 kilometers to access TB testing strongly preferred home-based testing.

### Willingness-to-Trade Simulations

In willingness-to-trade simulations, participants were on average, willing to trade 14.3% (range: 9.8-20.0% across countries) lower accuracy for the availability of rapid results (within 15 minutes) and 7.8% (range: 5.3-12.3%) lower accuracy for same-day results (**Table 2a**)^15^; participants across all countries were unwilling to trade a meaningful amount of accuracy for non-sputum sample types or non-health-facility-based testing. In willingness to pay analyses, on average, participants were willing to pay $2.90 out-of-pocket for the availability of rapid results (within 15 minutes) and $1.50 lower accuracy for same-day results, although this differed substantially by country with only participants in Vietnam and India being willing to incur any costs for rapid test availability (**Table 2b**)^15^.

**Table 2a.** Willingness to trade diagnostic accuracy for preferred TB test features overall and by country.

| Attribute/Level | Overall<br>(n=1033) | India<br>(n=200) | Philippines<br>(n=210) | South Africa<br>(n=215) | Uganda<br>(n=200) | Vietnam<br>(n=208) |
| --- | --- | --- | --- | --- | --- | --- |
| **Sample type** |  |  |  |  |  |  |
| Oral swab | 0% | 0% | 0.3% | 0% | 0% | 0.2% |
| Urine | 0% | 0% | 0% | 0% | 0% | 0% |
| Finger prick | 0% | 0% | 0% | 0% | 0% | 0% |
| Sputum | Ref. | Ref. | Ref. | Ref. | Ref. | Ref. |
| **Location of testing** |  |  |  |  |  |  |
| Home | 0% | 0% | 0% | 0% | 0% | 1.8% |
| Community | 0% | 0% | 0% | 0% | 0% | 0% |
| Hospital | Ref. | Ref. | Ref. | Ref. | Ref. | Ref. |
| **Time to result** |  |  |  |  |  |  |
| 15 minutes | 14.3% | 10.8% | 20%* | 19.8% | 20% | 9.8% |
| 3 hours | 7.8% | 6.0% | 10.4% | 10.0% | 12.3% | 5.3% |
| 1 day | Ref. | Ref. | Ref. | Ref. | Ref. | Ref. |

The willingness to trade analysis assumes that the cost of the TB test is free and initially has 90% sensitivity (based on Xpert Ultra for pulmonary TB)<sup>15</sup>. \*Willingness to trade was at least 20%, but the value is extrapolated beyond the range of what participants were directly asked to consider in the DCE.

**Table 2b.** Willingness to pay for preferred TB test features overall and by country.

| Attribute/Level | Overall<br>(n=1033) | India<br>(n=200) | Philippines<br>(n=210) | South Africa<br>(n=215) | Uganda<br>(n=200) | Vietnam<br>(n=208) |
| --- | --- | --- | --- | --- | --- | --- |
| **Sample type** |  |  |  |  |  |  |
| Oral swab | \$0 | \$0 | \$0.05 | \$0 | \$0 | \$0.10 |
| Urine | \$0 | \$0 | \$0 | \$0 | \$0 | \$0 |
| Finger prick | \$0 | \$0 | \$0 | \$0 | \$0 | \$0 |
| Sputum | Ref. | Ref. | Ref. | Ref. | Ref. | Ref. |
| **Location of testing** |  |  |  |  |  |  |
| Home | \$0 | \$0 | \$0 | \$0 | \$0 | \$1.25 |
| Community | \$0 | \$0 | \$0 | \$0 | \$0 | \$0 |
| Hospital | Ref. | Ref. | Ref. | Ref. | Ref. | Ref. |
| **Time to result** |  |  |  |  |  |  |
| 15 minutes | \$2.90 | \$3.20 | \$0 | \$0 | \$0 | \$4.00* |
| 3 hours | \$1.50 | \$1.65 | \$0 | \$0 | \$0 | \$3.70 |
| 1 day | Ref. | Ref. | Ref. | Ref. | Ref. | Ref. |

The willingness to pay analysis assumes that the cost of the TB test is free and initially has 90% sensitivity (based on Xpert Ultra for pulmonary TB)<sup>15</sup>. \*Willingness to pay was at least \$4.00 USD but the value is extrapolated beyond the range of what participants were directly asked to consider in the DCE.

## Discussion

In this multi-national study, we found that free cost, high diagnostic accuracy, and rapid results are the attributes that are universally valued by people undergoing evaluation for TB. The universal preference for high diagnostic accuracy aligns with the critical need for reliable TB tests to ensure accurate diagnosis, while the strong preference for free tests is expected given the socioeconomic challenges faced by many TB-affected people in high-burden countries.

There was a notable willingness among participants to trade 10-20% lower accuracy and among some to incur small out-of-pocket costs to access point-of-care tests that provide results within 15 minutes. This finding is aligned with a prior DCE among people with TB in Zambia^16^, and underscores the importance of timely health information as people undergoing TB testing may find it inconvenient and difficult to return on another day to the health facility for their TB test results due to incidental costs (e.g., lost wages, transport) and desire to initiate treatment as soon as possible, especially if feeling ill. Our results highlight the need for test developers and TPP guidelines to consider factors beyond high accuracy and adopt person-centered approaches that ensure TB diagnostic tools align with the preferences and practical needs of patients.

Across most countries, there was a strong preference for health-facility-based testing, although participants in Vietnam showed a preference for home-based testing. Community-based testing was the least preferred location. This is potentially limited by our study population, who were all enrolled at health facilities and had undergone facility-based testing in the prior three months. Health facilities may have been preferred despite being less convenient because people were already receiving care there and trust their health workers. Health centers may also be perceived as being more reliable, having better diagnostic equipment that ensures higher accuracy, providing immediate access to treatment, offering comprehensive care, and/or having experienced and knowledgeable healthcare professionals. Nonetheless, further research at lower-level health facilities and in community settings is needed to better understand the influence of location on TB test preferences and to assess whether similar preferences are observed in people who have not yet accessed care.

Non-sputum-based tests, such as point-of-care fingerpick blood tests, urine tests, and tongue-swab based tests, are key priorities of the current WHO TPP for TB tests^6^. However, our DCE results indicate that the sample type was the least important attribute for people accessing TB testing. Somewhat unexpectedly, sputum was the most preferred sample type overall. Among our participants, all of whom had recently undergone sputum testing, this preference may reflect greater familiarity and trust in its reliability for TB diagnosis, contributing to its selection over easier to collect sample types^17,18^. Notably, younger participants, those with prior TB, and individuals from Vietnam and the Philippines showed a preference for tongue swabs over sputum but were not willing to compromise on accuracy or pay more for these tests. These findings suggest that while there is some preference for easier to collect sample types, they are not a primary concern for people undergoing TB testing. This highlights the need for further research among populations who have not previously undergone sputum-based testing or are less likely to be able to produce sputum to better understand their preferences and potential acceptance of non-sputum-based diagnostics. Additionally, effective communication and education strategies may be required to increase public trust and acceptance of new non-sputum-based TB tests, which have always been the cornerstone of TB diagnosis.

The primary strength of this study is the large sample size that included participants from five diverse, high TB-burden countries in different geographical regions (sub-Saharan Africa, South Asia, and Southeast Asia), which improves the generalizability of our findings. Further, the DCE language, including attribute descriptions and the exercise overview, as well as representative graphics for different features, were refined and finalized with input from in-country partners and piloted in each country prior to enrollment to ensure local relevance and comprehension. Standardized procedures were used across sites to ensure comparability. Additionally, a participant education flipbook and props were shown to participants prior to the DCE to facilitate understanding. Ultimately, only 8.6% of participants were excluded for low-quality data that could indicate poor comprehension or inattention, and 94% of all participants included said the DCE was easy to understand.

Our study did, however, have some limitations. Foremost, despite our committed efforts to include participants representative of people who undergo TB testing in high-burden countries, our study only captured those who were already accessing TB testing services, rather than those indicative of the ‘missing millions’ who are not being reached by TB services. Thus, people with TB who can only be reached by community-based strategies may have preferences for TB testing that significantly differ from participants in our study. Future preference research should identify means for including harder-to-reach populations to ensure a more comprehensive understanding of preferences of TB affected communities. Furthermore, most participants had likely never provided non-sputum sample types or experienced non-health facility testing. While we tried to minimize this bias through education flipbooks and the use of props, because it is difficult for one to have informed preferences on something they have never experienced, this could have underestimated how much participants would value these features if available^19^.

In conclusion, our findings underscore the critical need for TB diagnostic tools that align with preferences of those undergoing testing, particularly prioritizing rapid results. While high diagnostic accuracy and minimal-to-no costs remain essential, the willingness of people to trade some accuracy and possibly pay for faster results highlights the value they place on timely health information. Currently, the preferences of people undergoing TB testing are absent from the WHO TPP, and future TPP updates and TB test developers should adopt person-centered approaches to ensure the practical needs of those undergoing TB testing are met. In particular, future TB research should focus on understanding the preferences of people affected by TB who have difficulty accessing facility-based TB testing.

## Data Availability

All data produced in the present study are included in the manuscript or available upon reasonable request to the authors

---

**Figure S1.**
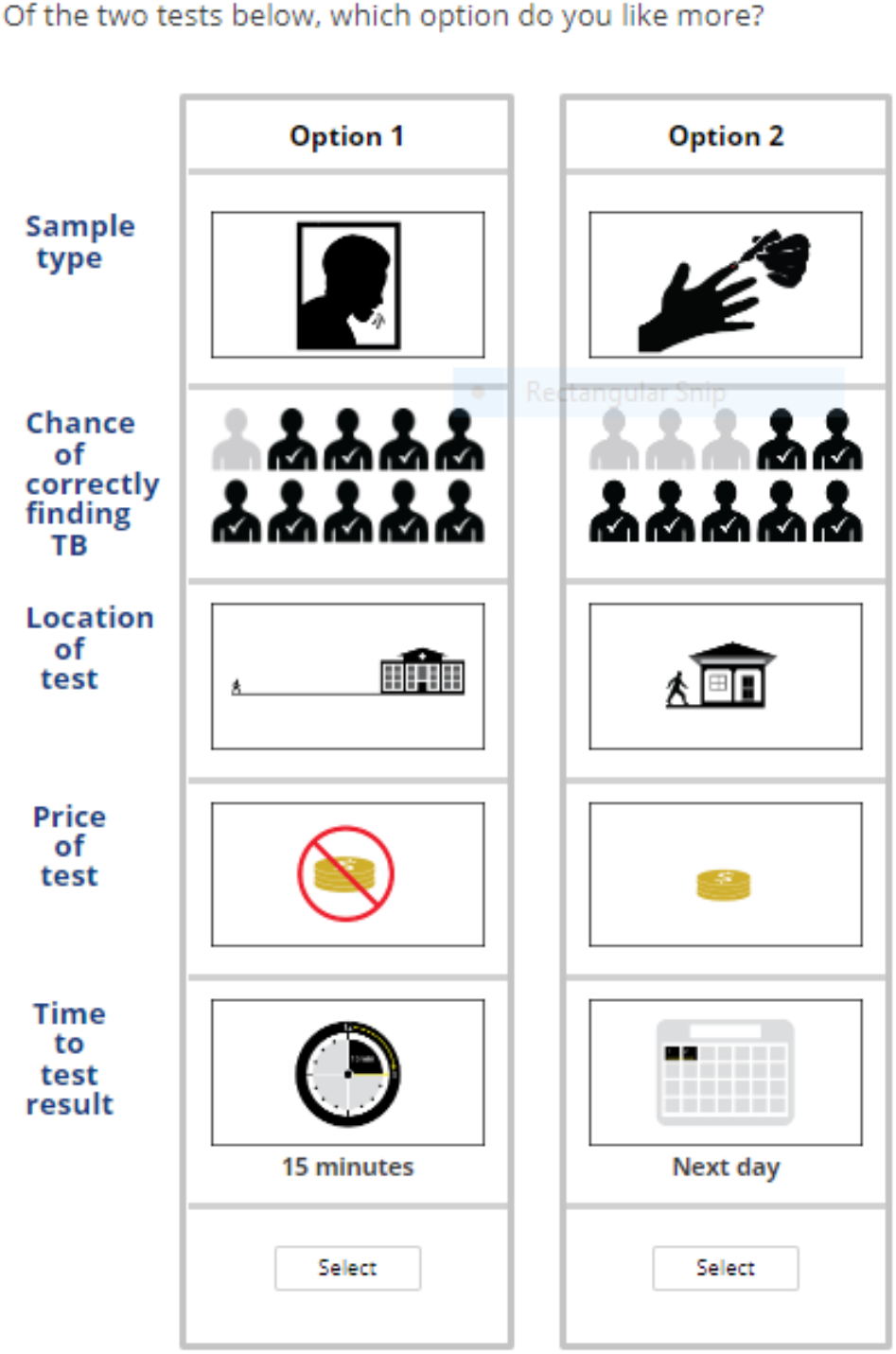
Example choice task as displayed to participants.

**Table S1.** Overview of country-specific settings, patient populations and presumptive TB definitions used for study inclusion.

| **Country** | **Study Setting** | **Patient Population** | **Average distance or time to facility** | **Presumptive TB definition** |
| --- | --- | --- | --- | --- |
| **India** | There are three locations:<br>DOTS clinic and pulmonary OPD at a large tertiary care hospital in Vellore, Tamil Nadu<br>Primary care clinic in local district of Vellore<br>Secondary care facility in a smaller city of Chittoor, Andhra Pradesh | At the large tertiary care hospital, patients are from up to 2000km away.<br>At the other two clinics, patients are generally local.<br>Disease severity generally ranges from mild to moderate at all locations. | 100 km radius; patients have a 10 minute to 2 hour travel time. | Symptom based screening: cough, fever, night sweats, weight loss for 2 or more weeks (any duration if PLHIV) |
| **Philippines** | TB DOTS clinic at a large hospital outside of the city of Manila | Patients come from surrounding districts. Disease severity generally ranges from mild to moderate. | 25 km radius; most patients have a 10 minute to 1 hour travel time | Symptom based screening: cough, fever, night sweats, weight loss for 2 or more weeks. |
| **South Africa** | Clinic located in smaller district of city of Cape Town | Patients are from the local district or may come from neighboring districts. Disease severity generally ranges from mild to moderate. Any severe cases are referred to a nearby hospital. | Most patients are a 5-10 minute walk from the clinic. | Symptom based screening: cough, fever, night sweats, weight loss for 2 or more weeks.<br>OR<br>Patients who are:<br>on ART, newly diagnosed with HIV,<br>pregnant with HIV,<br>anyone who received TB treatment in the past two years, close contacts. |
| **Uganda** | There are two locations:<br>National referral hospital located on the outside of city of Kampala<br>City hospital located in the city center of Kampala | Patients come from districts within Kampala and some neighboring districts and villages.<br>Patients come from the urban areas of Kampala as well as non-residents who commute to Kampala for work.<br>Disease severity generally ranges from mild to severe. | 50 km radius, most patients have a 5 minute to 2 hour travel time. | Symptom based screening: cough, fever, night sweats, or weight loss for 2 or more weeks. |
| Vietnam | Lung hospital in city of Hanoi | Majority of patients come from within the city, a small proportion from outside the city. Disease severity generally ranges from mild to moderate. | 10-15 km radius; most patients have <1 hour travel time | Symptom based screening: cough, fever, night sweats, weight loss for 2 or more weeks.<br>OR<br>TB risk factors such as HIV, close contact of person with TB, substance use, chronic disease, immunosuppressant use if symptomatic or asymptomatic. |

**Table S2.** Sensitivity analyses: Relative importances and mean preference weights.

|  | **Overall**<br>**(n=1,129)** | **India**<br>**(n=236)** | **Philippines**<br>**(n=244)** | **South Africa**<br>**(n=233)** | **Uganda**<br>**(n=205)** | **Vietnam**<br>**(n=211)** |
| --- | --- | --- | --- | --- | --- | --- |
| **Sample type** | 13.3 | 15.4 | 14.5 | 13.3 | 11.1 | 14.5 |
| **Sputum** | 3.41 | 1.12 | -0.38 | 4.26 | 6.04 | 6.85 |
| **Oral swab** | 3.26 | 2.40 | 2.89 | 1.68 | 3.35 | 6.34 |
| **Urine** | -2.91 | -0.78 | -1.20 | -4.23 | -3.22 | -5.52 |
| **Fingerprick** | -3.76 | -2.72 | -1.32 | -1.71 | -6.17 | -7.67 |
| **Accuracy** | 25.2 | 27.1 | 19.2 | 21.5 | 20.4 | 19.2 |
| **70%** | -55.04 | -59.13 | -36.34 | -45.90 | -43.0 | 96.65 |
| **80%** | 0.57 | 0.65 | 1.70 | 1.80 | 1.17 | -2.75 |
| **90%** | 54.47 | 58.50 | 34.64 | 44.10 | 41.82 | -93.91 |
| **Location** | 18.0 | 17.9 | 19.8 | 14.9 | 24.1 | 19.8 |
| **Home** | -17.54 | -11.82 | -30.02 | -10.81 | -47.61 | 12.26 |
| **Community location** | -4.16 | -4.59 | -3.89 | -3.62 | -0.66 | -7.97 |
| **Health facility** | 21.70 | 16.41 | 33.91 | 14.43 | 48.28 | -4.28 |
| **Cost** | 25.4 | 22.6 | 29.5 | 31.9 | 27.2 | 29.5 |
| **Free** | 55.81 | 43.44 | 68.53 | 68.93 | 65.46 | 31.10 |
| **\$2** | -2.10 | -2.16 | -4.06 | -0.76 | -1.44 | -1.90 |
| **\$4** | -53.71 | -41.30 | -64.47 | -68.17 | -64.01 | -29.19 |
| **Time** | 18.2 | 17.0 | 17.1 | 18.4 | 17.2 | 17.1 |
| **15 minutes** | 37.34 | 32.90 | 30.14 | 29.24 | 37.73 | 48.22 |
| **3 hours** | 4.07 | 4.31 | 3.10 | 2.84 | 4.97 | 5.43 |
| **Next day** | -41.42 | -37.20 | -33.25 | -42.08 | -42.70 | -53.64 |

## References

1. World Health Organization. *Global Tuberculosis Report* 2023. World Health Organization; 2023. https://iris.who.int/bitstream/handle/10665/373828/9789240083851-eng.pdf?sequence=1

2. Storla D, Yimer S, Bjune G. A systematic review of delay in the diagnosis and treatment of tuberculosis. BMC Public Health. 2008;8(15). doi:10.1186/1471-2458-8-15

3. Myburgh H, Baloyi D, Loveday M, et al. A scoping review of patient-centred tuberculosis care interventions: Gaps and opportunities. PLoS Glob Public Health. 2023;3(2). 10.1371/journal.pgph.0001357

4. World Health Organization. People-Centred Framework for Tuberculosis Programme Planning and Prioritization. World Health Organization; 2019.

5. Kik SV, Denkinger CM, Casenghi M, Vadnais C, Pai M. Tuberculosis diagnostics: which target product profiles should be prioritised? Eur Respir J. 2014;44(2):537–540. doi:10.1183/09031936.00027714

6. World Health Organization. High Priority Target Product Profiles for New Tuberculosis Diagnostics: Report of a Consensus Meeting. World Health Organization; 2014. https://www.who.int/publications/i/item/WHO-HTM-TB-2014.18

7. Kerkhoff A, West N, del mar Castro M, et al. Placing the values and preferences of people most affected by TB at the center of screening and testing: an approach for reaching the unreached. BMC Glob Public Health. 2023;1(27). 10.1186/s44263-023-00027-0

8. Our Big Mac index shows how burger prices differ across borders. The Economist. Published online January 2024. https://www.economist.com/big-mac-index

9. Menapace L, Raffaelli R. Unraveling hypothetical bias in discrete choice experiments. J Econ Behav Organ. 2020;176:416–430.

10. Orme B. Getting Started with Conjoint Analysis: Strategies for Product Design and Pricing Research. Research Publishers LLC. Research Publishers LLC; 2010.

11. Orme B, Chrzan K. Becoming an Expert in Conjoint Analysis. Research Publishers LLC; 2017.

12. Johnson F, Yang J, Reed S. The Internal Validity of Discrete Choice Experiment Data: A Testing Tool for Quantitative Assessments. Value Health. 2019;22(2). 10.1016/j.jval.2018.07.876

13. Orme B. Consistency Cutoffs to Identify “Bad” Respondents in CBC, ACBC, and MaxDiff. Sawtooth Software; 2019. https://ssistaging.com/resources/technical-papers/consistency-cutoffs-to-identify-bad-respondents-in-cbc-acbc-and-maxdiff

14. Kremser M. How to Improve Your Survey Data Quality. Sawtooth Software. Published October 24, 2022. https://sawtoothsoftware.com/resources/blog/posts/Improve-Survey-Data-Quality-With-Root-Likelihood

15. Zifodya JS, Kreniske JS, Schiller I, et al. Xpert Ultra versus Xpert MTB/RIF for pulmonary tuberculosis and rifampicin resistance in adults with presumptive pulmonary tuberculosis. Cochrane Infectious Diseases Group, ed. Cochrane Database Syst Rev. 2021;2021(5). doi:10.1002/14651858.CD009593.pub5

16. Kerkhoff A, Kagujje M, Nyangu S, et al. Pathways to care and preferences for improving tuberculosis services among tuberculosis patients in Zambia: A discrete choice experiment. PloS One. 2021;16(8). doi:doi: 10.1371/journal.pone.0252095

17. Bustamante G, Liebermann E, McNair K, Fontenot HB. Women’s perceptions and preferences for cervical cancer screening in light of updated guidelines. J Am Assoc Nurse Pract. 2023;35(11):699–707. doi:10.1097/JXX.0000000000000923

18. Lama Y, Qin Y, Nan X, et al. Human Papillomavirus Vaccine Acceptability and Campaign Message Preferences Among African American Parents: a Qualitative Study. J Cancer Educ. 2022;37(6):1691–1701. doi:10.1007/s13187-021-02014-1

19. Byrne RL, Wingfield T, Adams ER, et al. Finding the missed millions: innovations to bring tuberculosis diagnosis closer to key populations. BMC Glob Public Health. 2024;2(1):33. doi:10.1186/s44263-024-00063-4

