## Appendix 1 for "A Multi-Country Evaluation of Patient Preferences for Tuberculosis Diagnostics: A Discrete Choice Experiment to Inform WHO Target Product Profiles"

UCSF

R2D2 TB Network

### R2D2 DCE Patient Education Booklet

As mentioned before, we are gathering information from people like you who have recently been tested for TB, to understand their preferences for TB testing. For example, what features of a TB test are the most important and appealing  to you? The information from the study will help us develop tests for TB that are better aligned with what people want.

Before we begin, we would like to make sure that you understand the survey. The options for tests that we ask you to give your opinion on are hypothetical, which means that many of these tests do not yet exist. It also means that your responses will not affect your current TB treatment plan or overall healthcare in any way.

As part of this survey, we will ask you to consider your preferences for 5 different TB test features. I’d like to start by reviewing those features.

- The first option for a new tuberculosis test is the sample type that you provide and that is tested for tuberculosis.
- We will ask you about four types of samples that you would prefer to provide for tuberculosis testing.

### Sample types

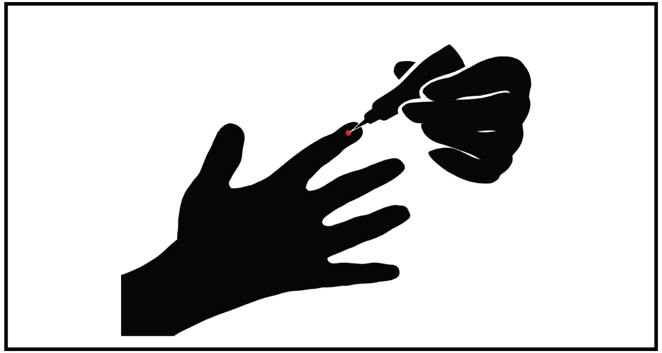

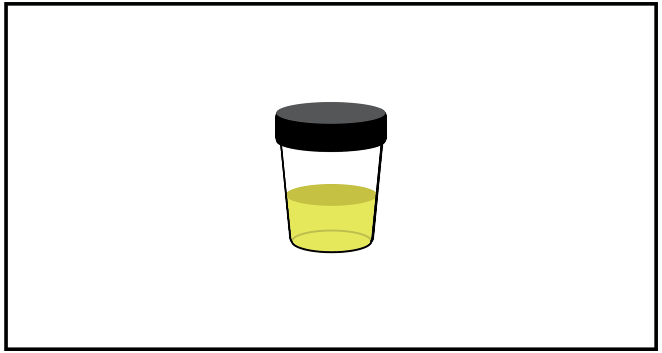

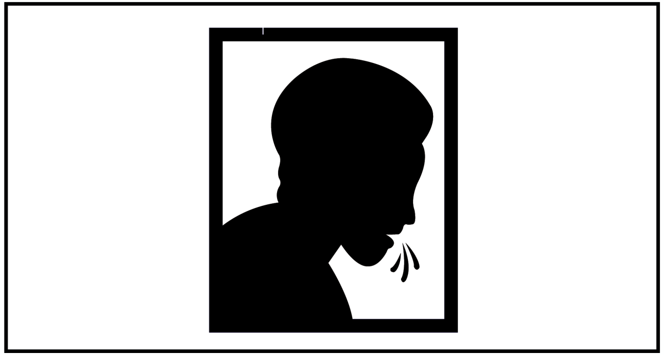

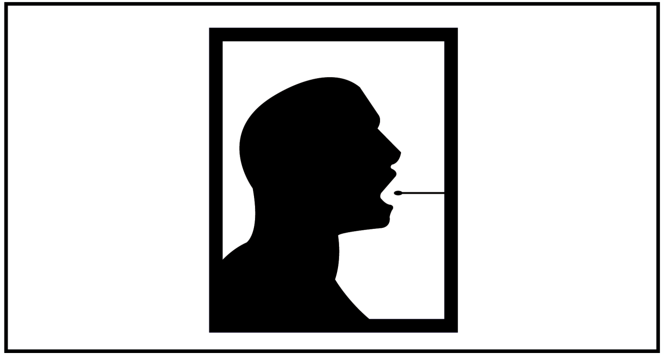

- The first sample type is sputum. This requires you to take deep breaths and cough hard until you feel a thick mucous in the back of your throat. This sample is then spit into a cup. This option looks like the graphic on the front of this card.

- I will now demonstrate this process using this collection cup.

### Sample type: Sputum

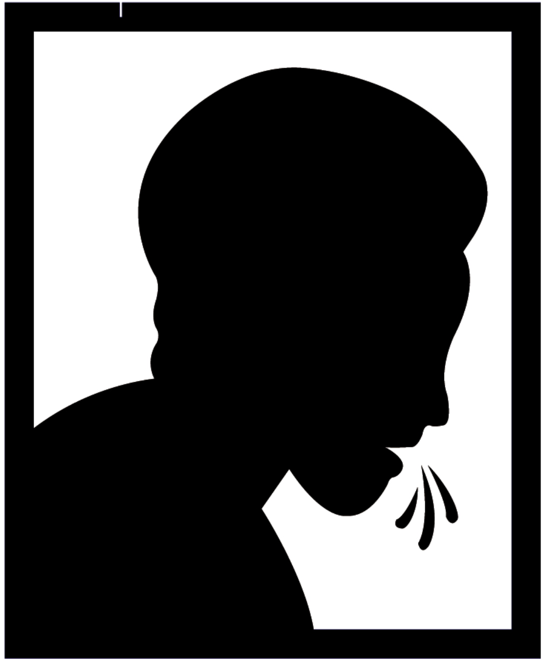

- The second sample type is a blood finger prick. This requires using a very small, sharp blade to obtain a very small amount of blood from your finger.
- This option looks like the graphic on the front of this card.
- Here is an example of this device. You put it close to your finger like this and push the button to draw a drop of blood for the test.”

### Sample type: Finger prick

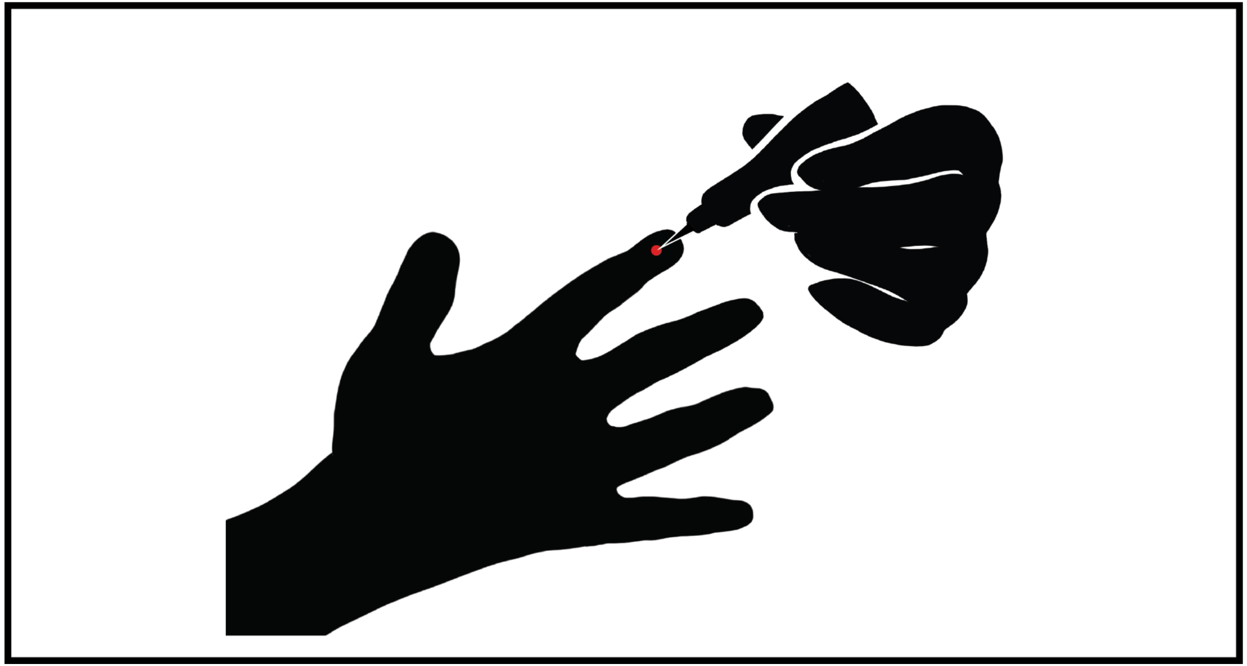

- The third sample type is urine. This requires you to start peeing and then collect a small amount of pee in a plastic cup.
- This option looks like the graphic on the front of this card.
- This is what the collection cup looks like that is used for this test option.

### Sample type: Urine

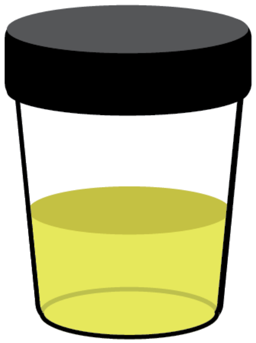

- The fourth sample type is a tongue swab. This requires you to stick out your tongue so that it can be swabbed for about 15 seconds. This option looks like the graphic on the front of this card.

- I will now demonstrate this process using this swab.

### Sample type: Tongue swab

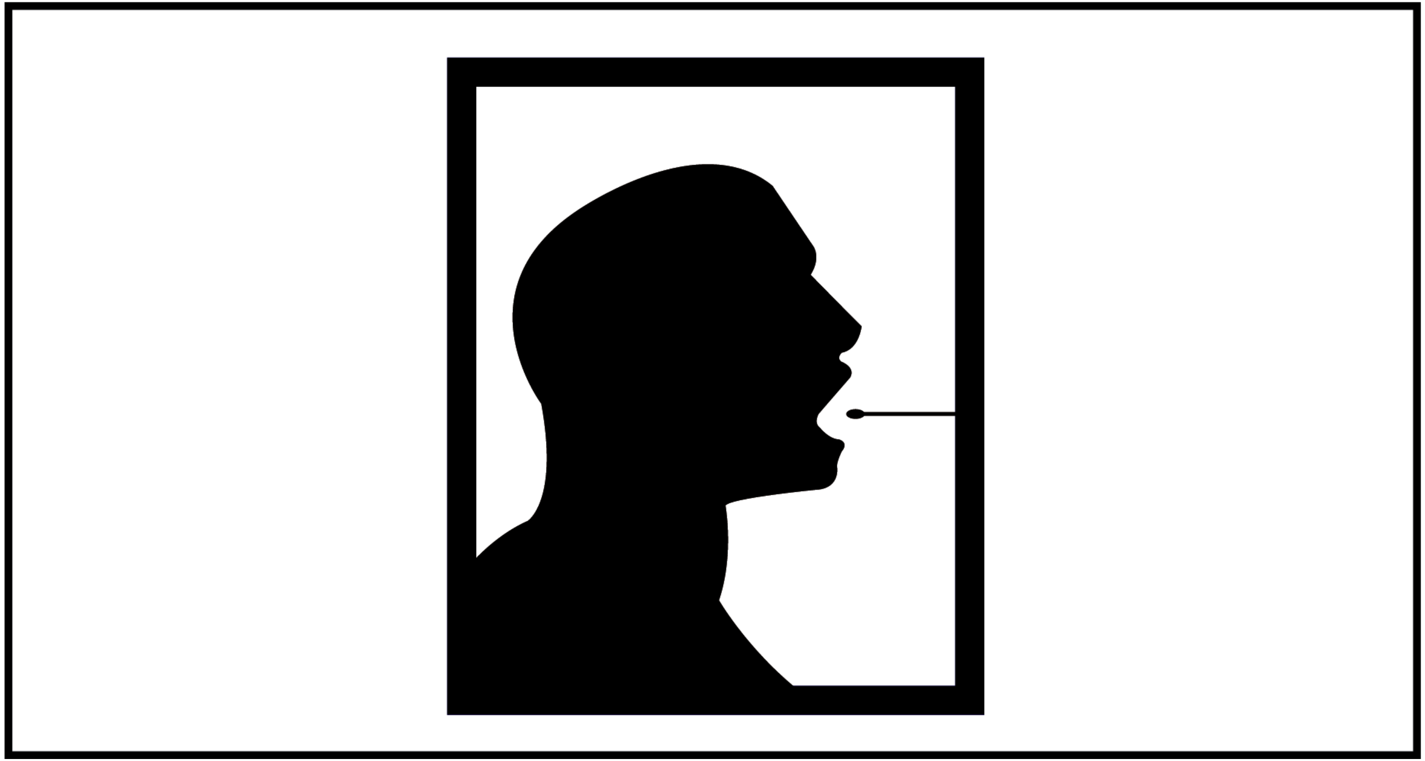

- Do you have any questions about what these pictures mean?

### Sample types

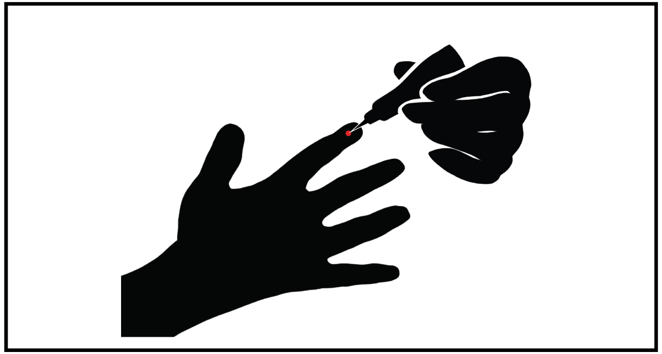

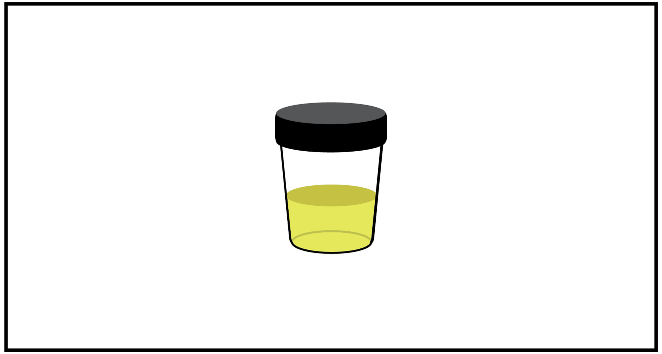

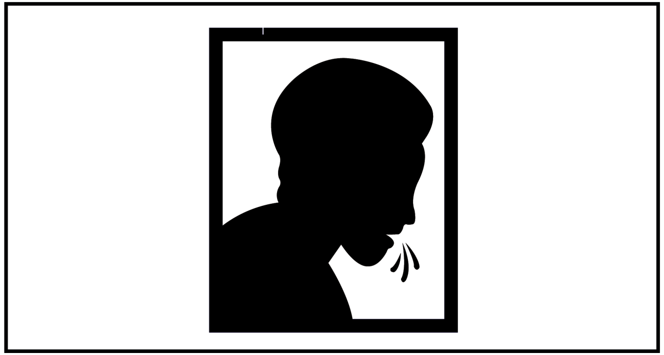

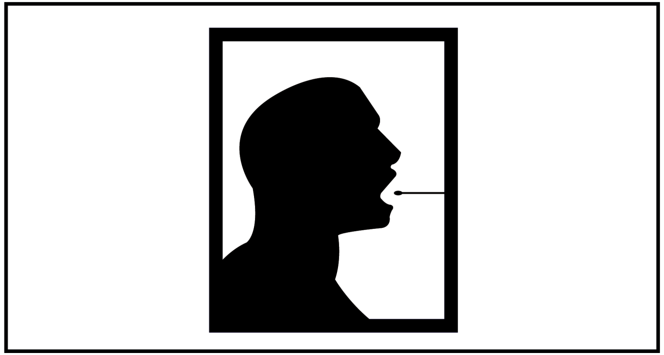

- The second option for a new tuberculosis test is how well the test performs at detecting tuberculosis, also known as accuracy.

- We will ask you about three different levels of accuracy that you would prefer for a tuberculosis test.

### Accuracy

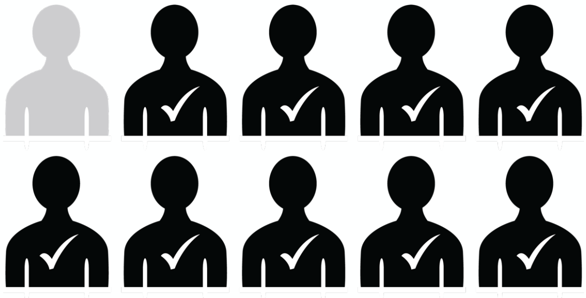

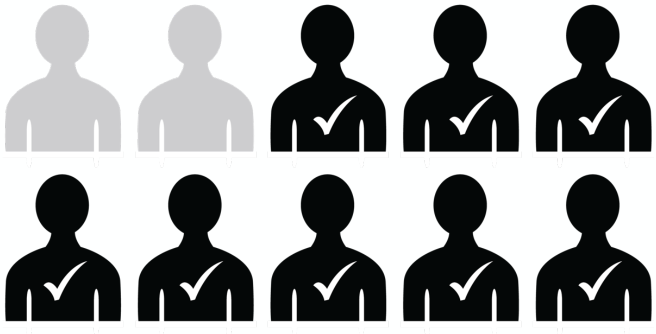

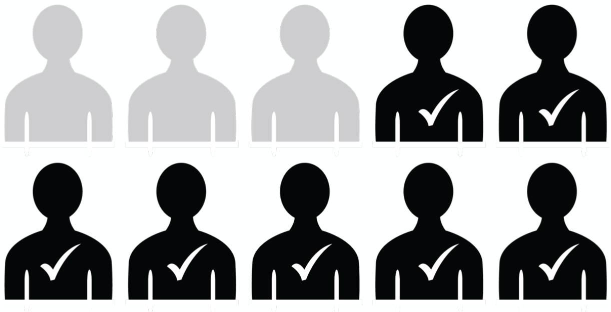

- The chance that the test misses tuberculosis when you have tuberculosis is: 1 in 10. This means that if 10 people had tuberculosis disease, the test would correctly detect tuberculosis in 9 persons, but would miss tuberculosis in 1 person. This option looks like the graphic on the front of this card.

### Accuracy: 9 in 10

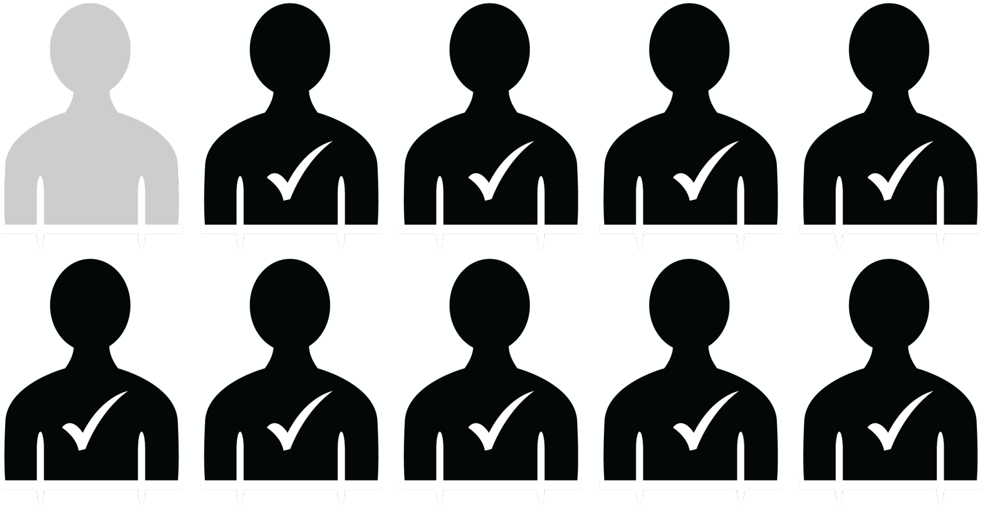

- The chance that the test misses tuberculosis when you have tuberculosis is: 2 in 10. This means that if 10 people had tuberculosis disease, the test would correctly detect tuberculosis in 8 persons, but would miss tuberculosis in 2 persons.
- This option looks like the graphic on the front of this card.

### Accuracy: 8 in 10

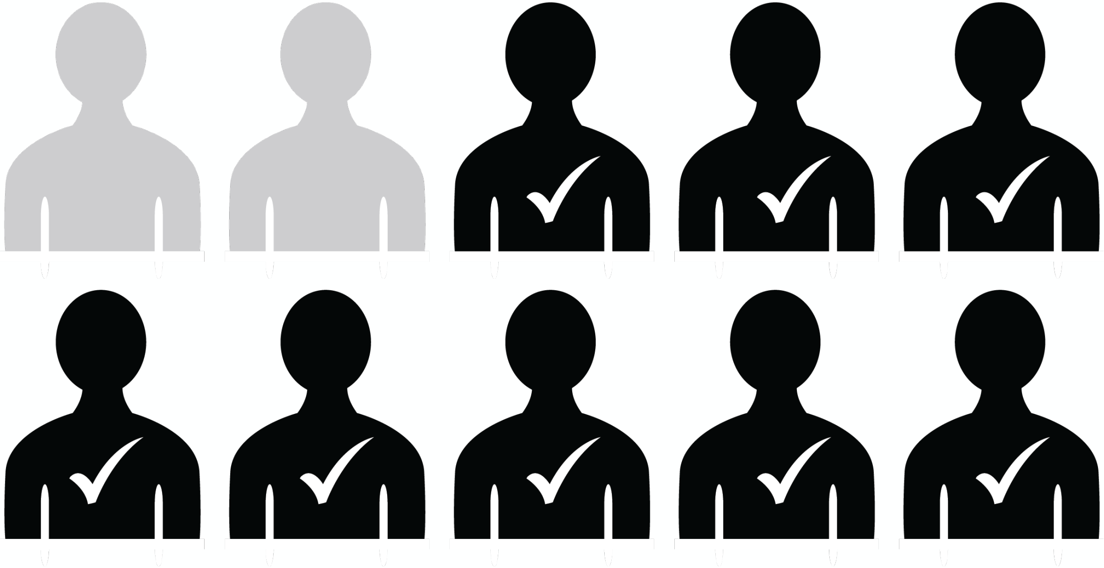

- The chance that the test misses tuberculosis when you have tuberculosis is: 3 in 10. This means that if 10 people had tuberculosis disease, the test would correctly detect tuberculosis in 7 persons, but would miss tuberculosis in 3 persons.
- This option looks like the graphic on the front of this card.

### Accuracy: 7 in 10

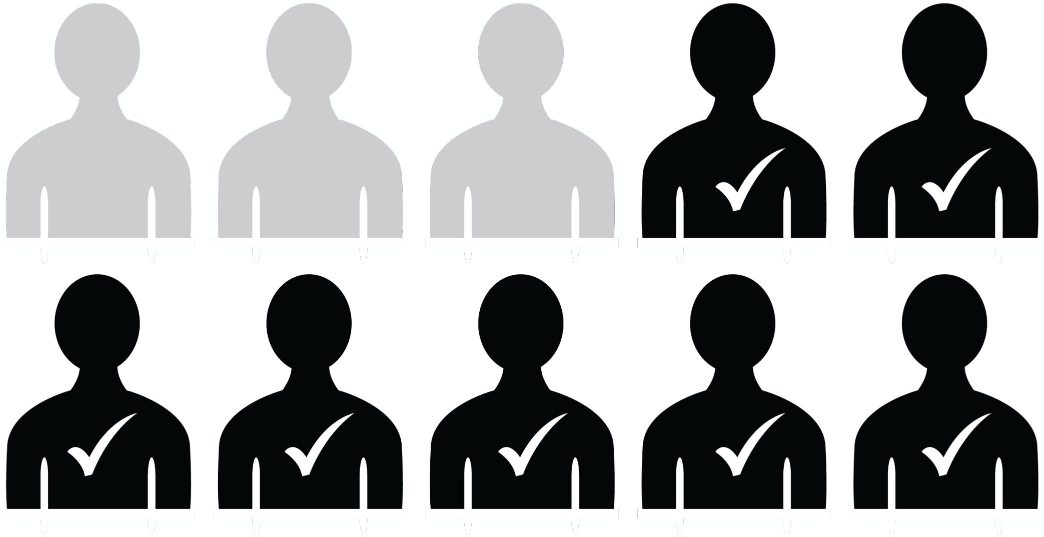

- Do you have any questions about what these pictures mean?

### Accuracy

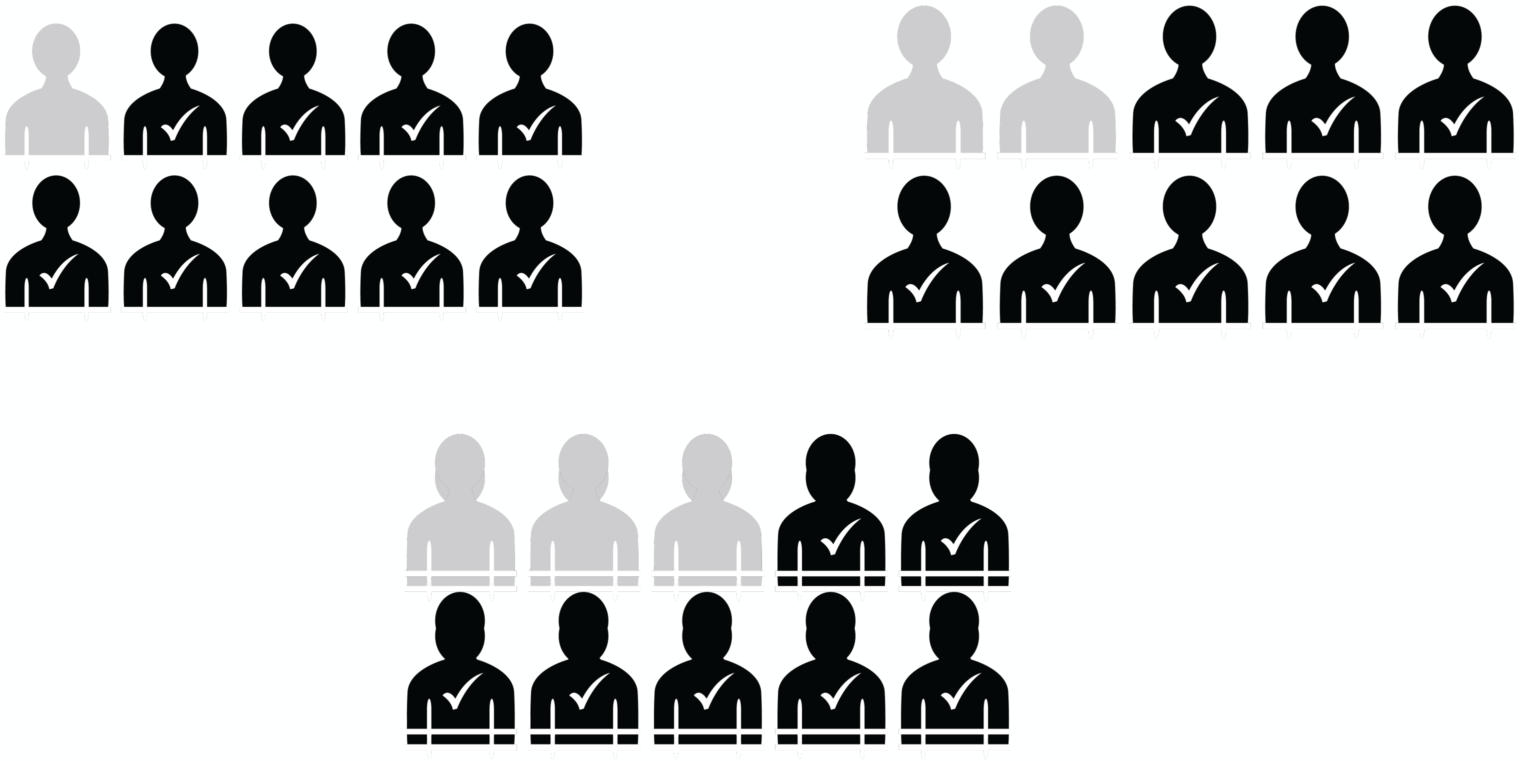

- What does this option mean?

  Participant should say some variation of: this test would find 7 out of 10 TB cases, this test would miss 3 of 10 TB positive cases.

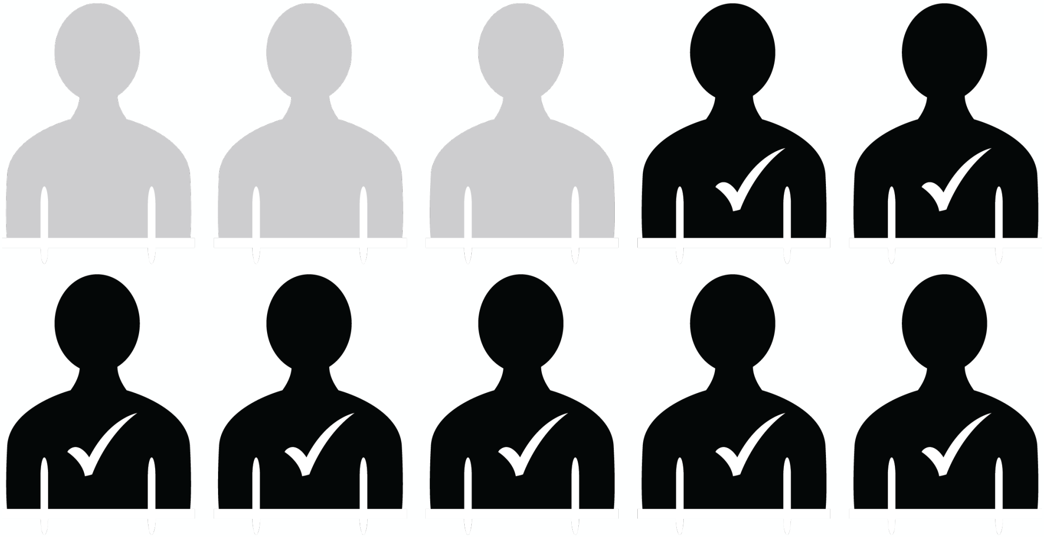

- The third option for a new tuberculosis test is the location where the tuberculosis test is performed.

- We will ask you about three different locations where you would prefer to undertake tuberculosis testing.

### Location of testing

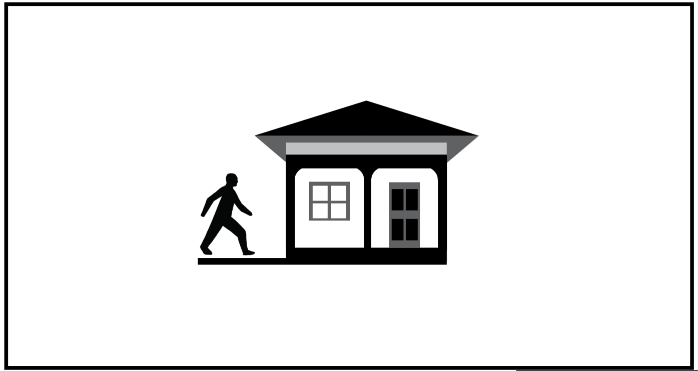

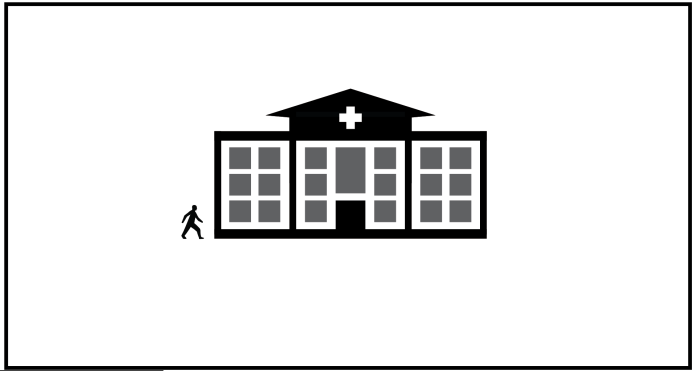

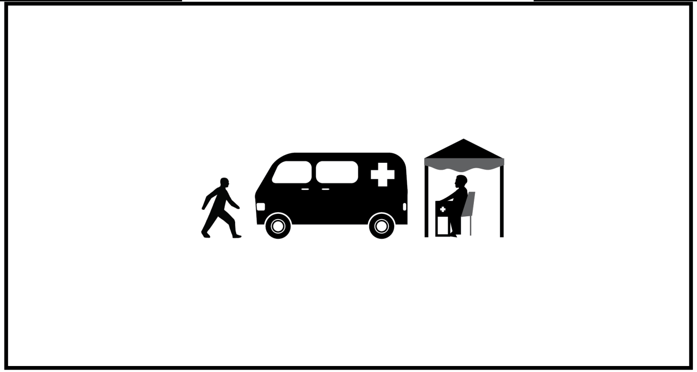

- The first location is your home. This means that a tuberculosis test could performed in your home. You must first get the test from elsewhere such as a pharmacy, shop, or stand in the community, but it can be performed at home at any time after you get the test.
- You can perform the test by yourself, or a trusted friend or family member can help you.

### Location: Home test

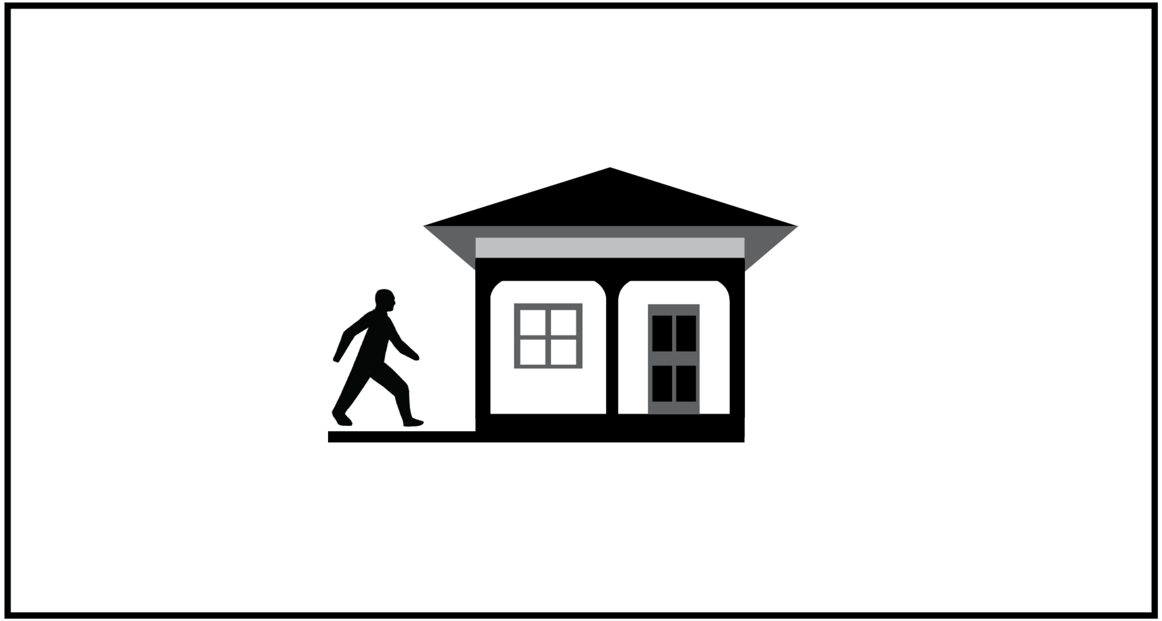

- The second location is a community site. This means that a tuberculosis test can be performed at a convenient location in your community, such as a smaller health clinic or pharmacy, or another community location that conducts health testing. This community location would not necessarily be affiliated with the nearest hospital or larger health facility and would likely be closer to your home than the nearest health facility.
- You must travel to the community site to get tested.

### Location: Community site

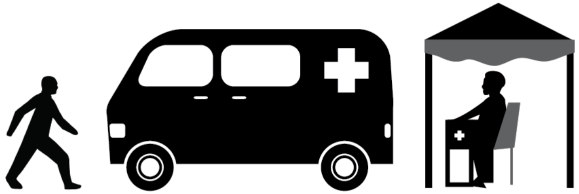

The third location is a health facility. This means that a tuberculosis test can only be performed at a health facility or a hospital, like this one. You must travel to the health facility to get tested.

### Location: Hospital

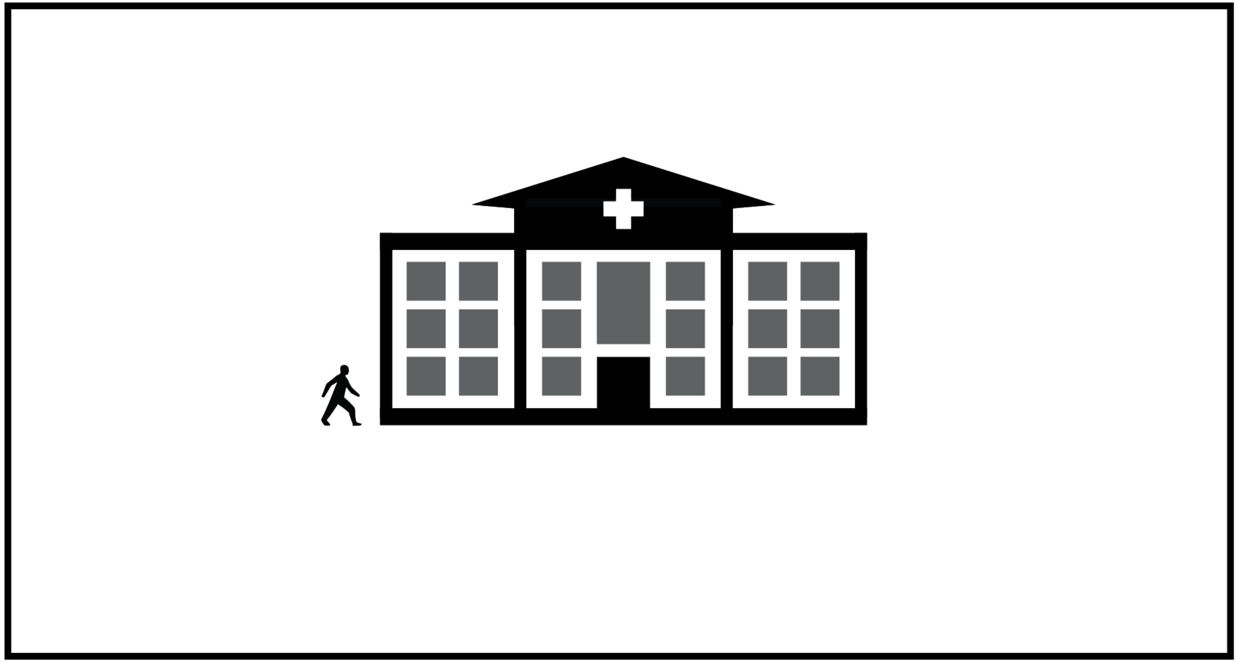

- Do you have any questions about these pictures and what they represent?

### Location of testing

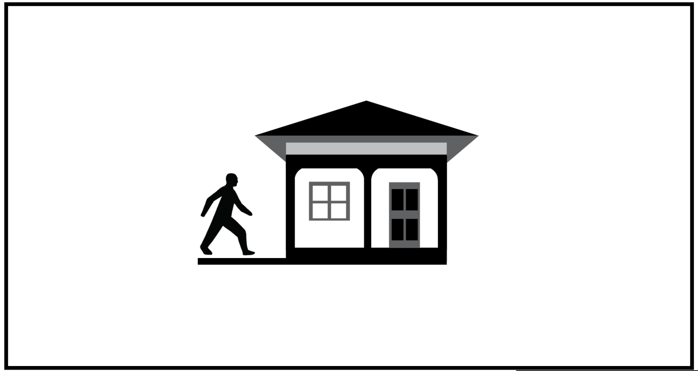

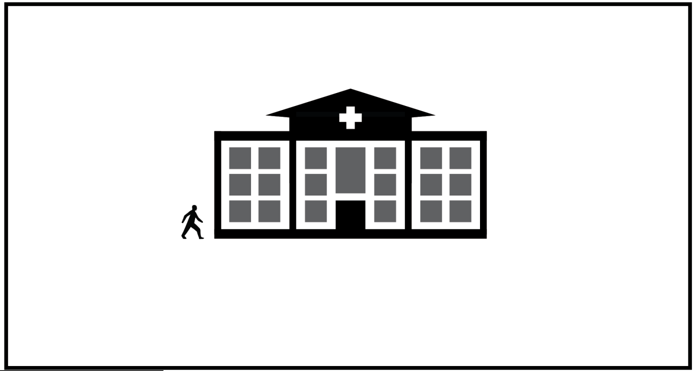

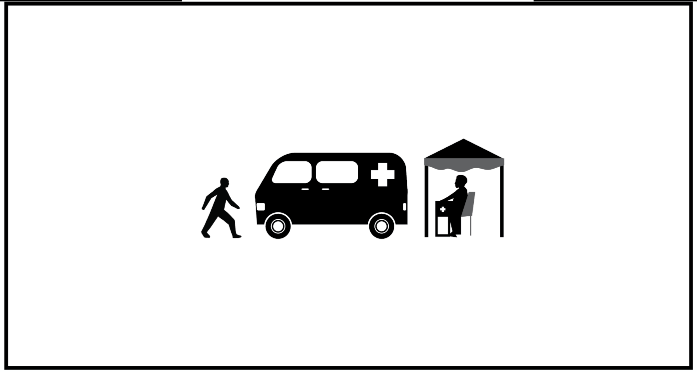

- The fourth option for a new tuberculosis test is the time to results of your test. This is the amount of time it takes to receive the results of your tuberculosis test after you have provided a sample.
- We will ask you about three different durations that you would prefer for how long you need to wait for your test results.

### Time to test result

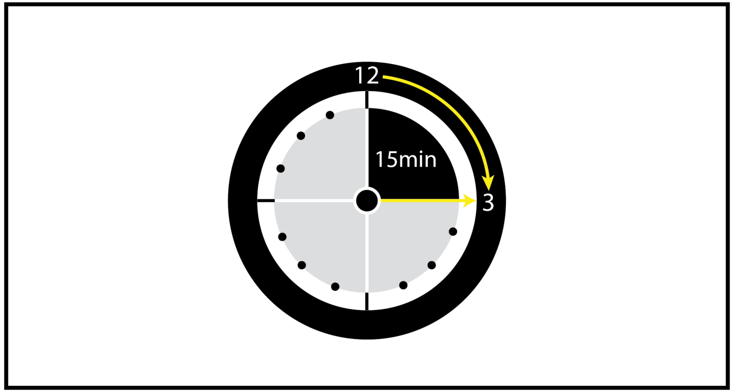


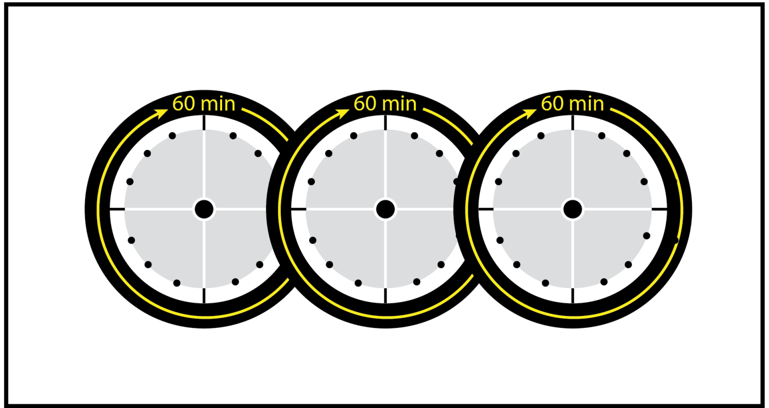


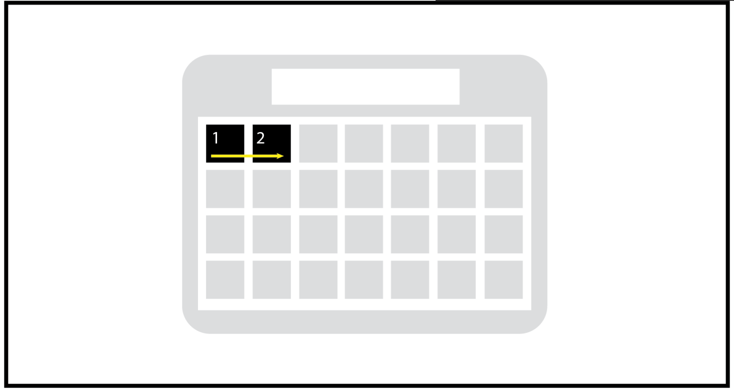

- The first amount of time is 15 minutes. This means that after you provide a sample, the results of your tuberculosis test will be available in 15 minutes.

### Time to test result: 15 minutes

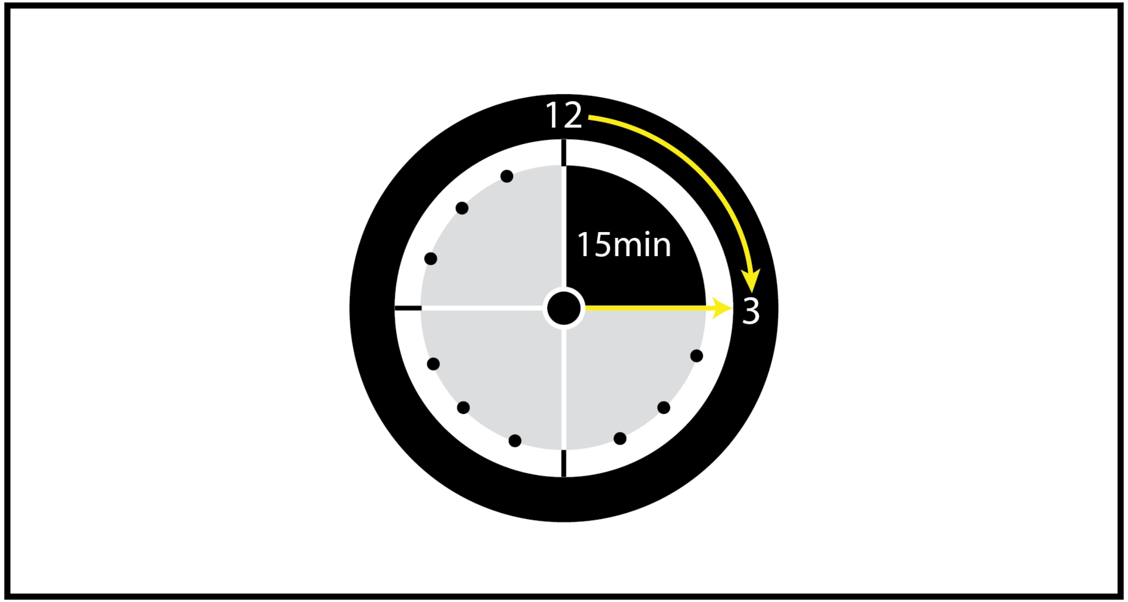

- The second amount of time is 3 hours. This means that after you provide a sample, the results of your tuberculosis test will be available in 3 hours.

### Time to test result: 3 hours

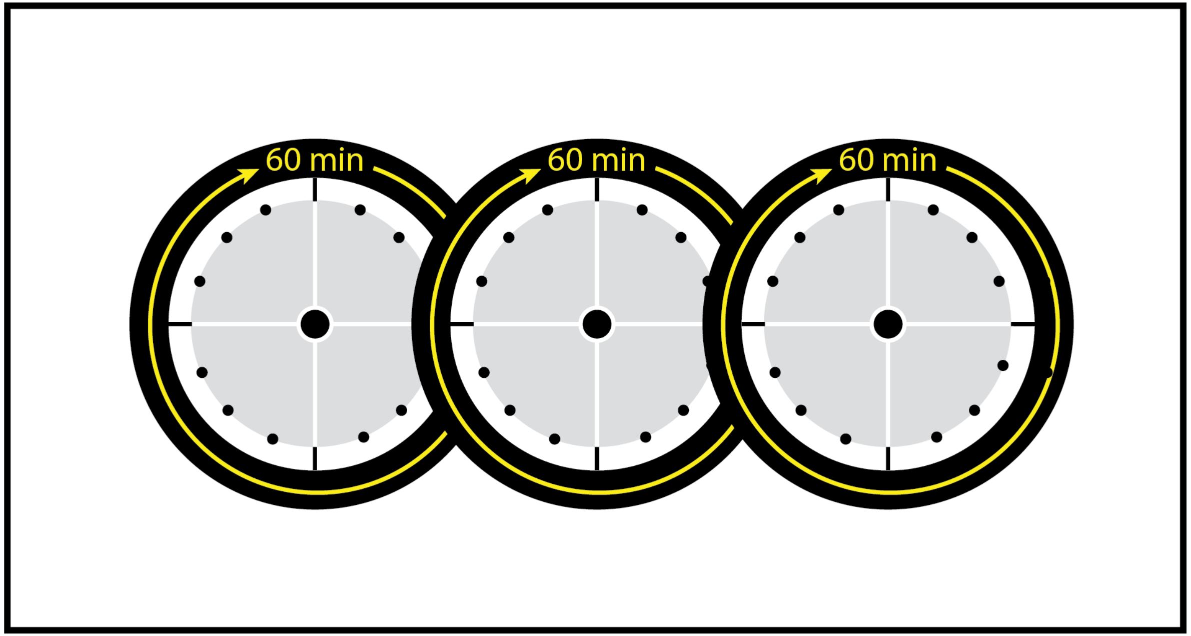

- The third amount of time is next day. This means that after you provide a sample, the results of your tuberculosis test won’t be available until the next day.

### Time to test result: Next day

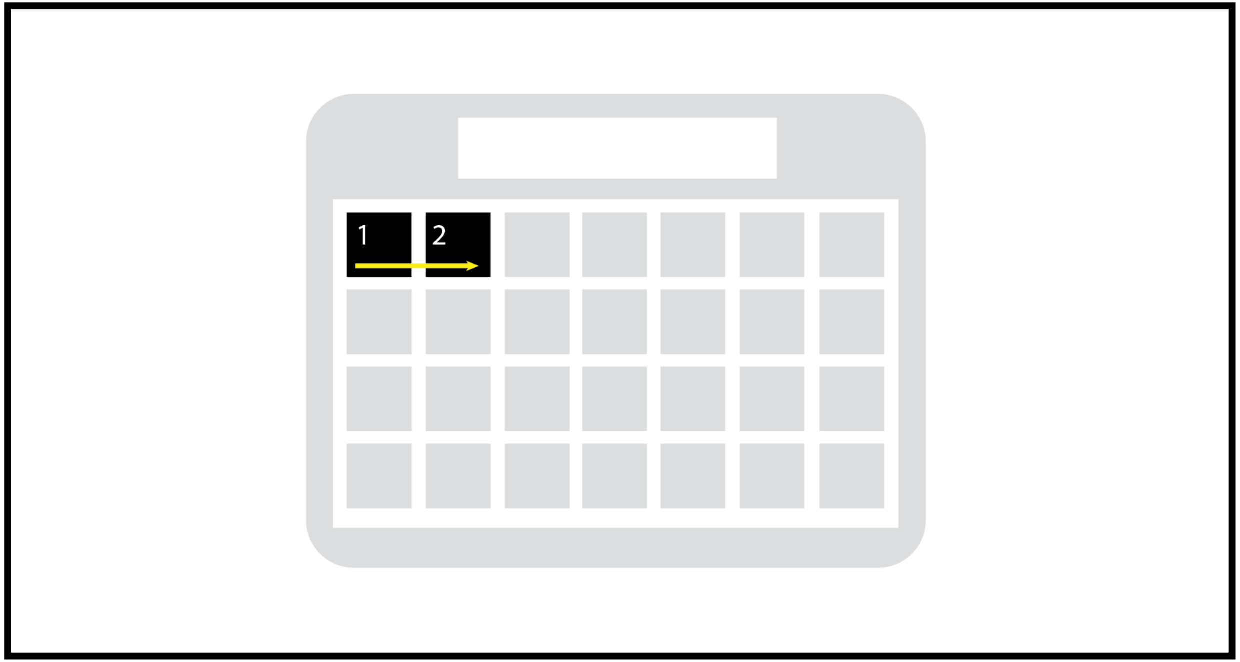

- Do you have any questions about what each picture represents?

### Time to test result

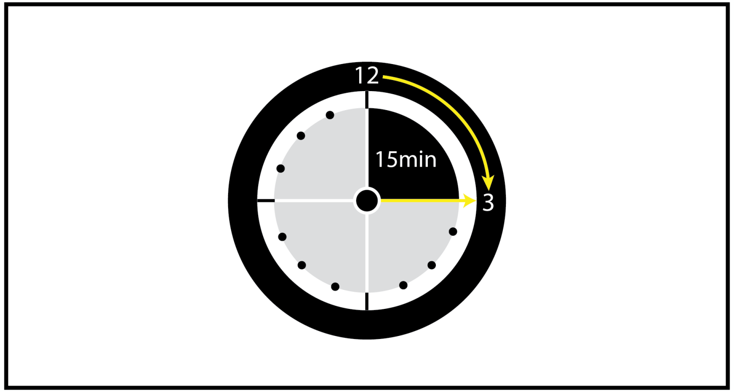


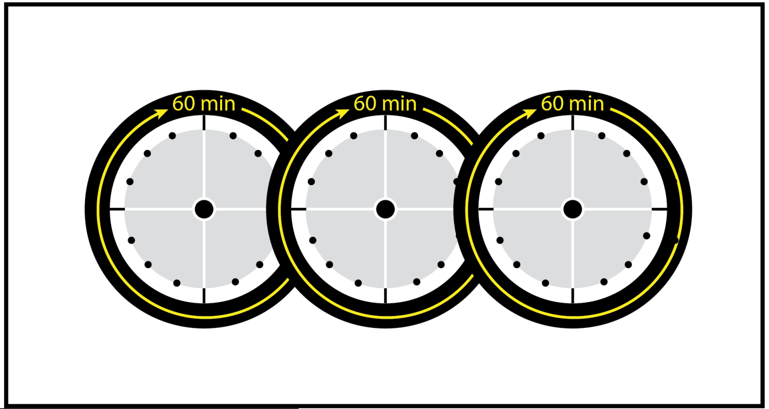


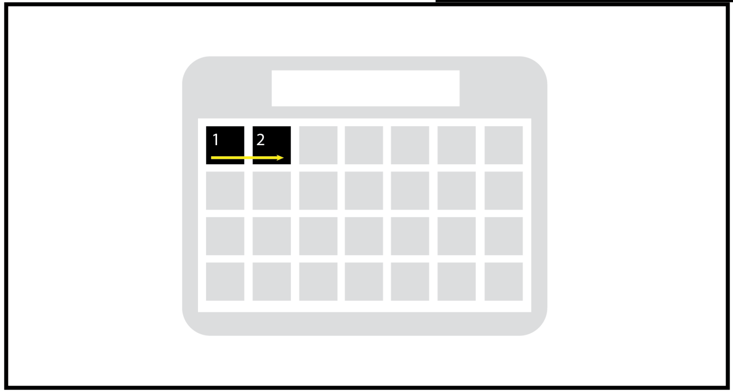

- The fifth option is the cost you need to pay for the test. This is the cost of the test itself that you would need to pay in addition to any costs for travel or time off from work.

- We will ask you about three different costs that you might pay for a tuberculosis test.

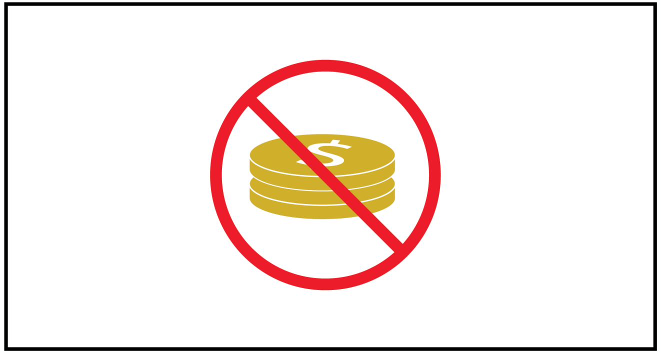

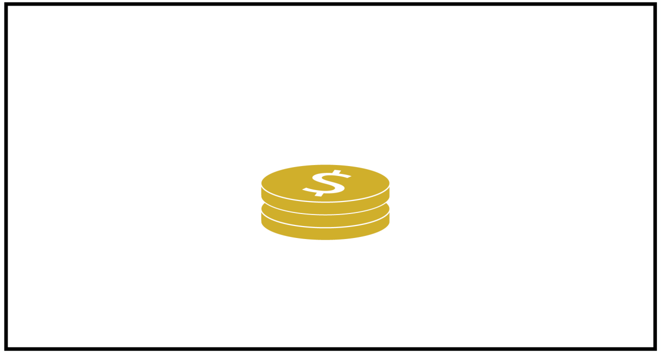

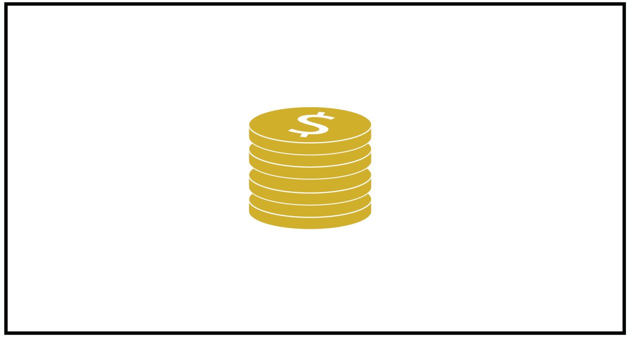

- The first amount is **FREE**. This means you would not pay any amount to access a tuberculosis test.

### Cost of test: Free

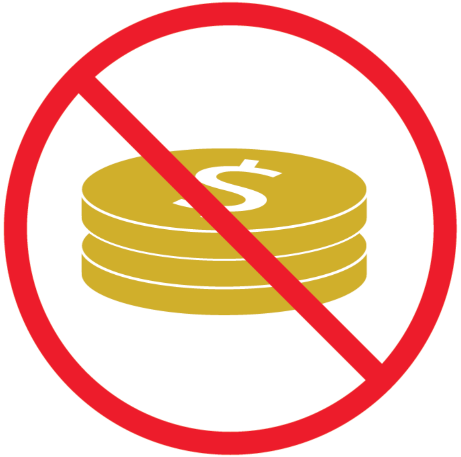

- The next amount is **$2 (country equivalent)** This means you would pay **$2** to access a tuberculosis test.

### Cost of test: $2

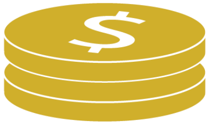

- The next amount is **$4 (country equivalent)**. This means you would pay **$4** to access a tuberculosis test.

### Cost of test: $4

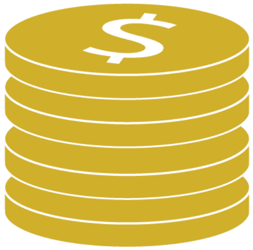

- Do you have any questions about the pictures shown to you?

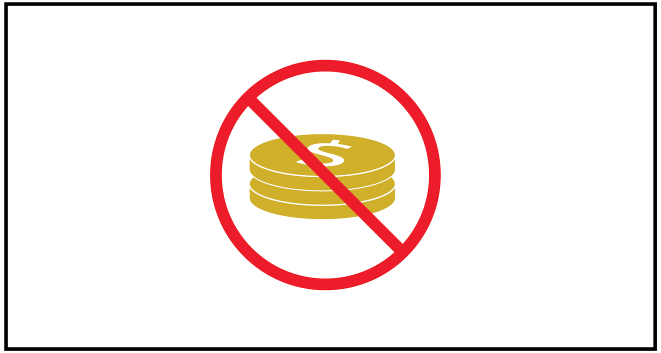

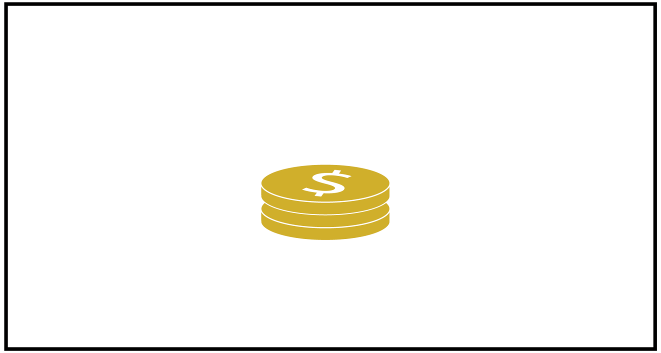

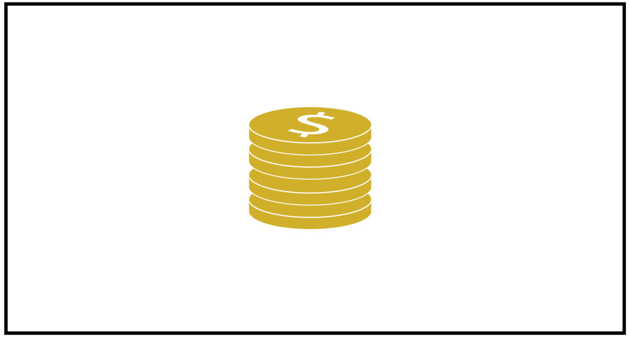

- I will now show you an example question of the activity on the study tablet/computer.
